# Kauro, a graph-based chatbot for high-fidelity information transmission conversations

**DOI:** 10.64898/2026.01.30.25342358

**Authors:** Charles Hadley King, Rebekah Barrick, Miguel Almalvez, Kirsten Blanco, Ivan De Dios, Vincent A. Fusaro, Emmanuèle Délot, Chris Donohue, Seth Berger, Changrui Xiao, UCI GREGoR Site, Eric Vilain, Jonathan LoTempio

## Abstract

Across biomedical research and care, many conversations transmit information with profound practical, ethical, and legal consequences. The process of informed consent, where individuals decide to join a study or accept clinical care, is perhaps the most consequential, yet it is also complex, labor-intensive, and variable across sites. Existing platforms for information transmission in the informed consent context largely reproduce static documents and lack reproducibility or auditability, while generative chatbots offer flexibility at the cost of stochasticity, hallucination, and regulatory risk. We present Kauro, an open-source, graph-based chatbot that encodes scripted conversations as version-controlled JavaScript Object Notation (JSON) structures, enabling deterministic traversal (ie, paths through the graph), complete audit logging, and IRB-verifiable oversight. Its modular separation of client, server, and script ensures portability across institutions. By operationalizing constraint rather than flexibility, Kauro reframes deployment of machine intelligence in biomedical communication with reproducibility and auditability, offering a scalable platform generalizable to any domain where conversations demand safety, precision, and trust.

## Introduction

There are many conversations that take place in the biomedical research and medical care context where critical information is transmitted between parties^1–3^. The first, and perhaps most important conversation is the informed consent conversation, where a person agrees to become a participant in a study, or to receive care^4^. Informed consent is the legally-regulated cornerstone of ethical biomedical research^5^. The consent conversation is complicated by the fact that the benefits and risks of participation can last for many years, or a person’s life and can impact the biological family and cultural cohorts of an individual participant (or indeed patient)^6–9^.

In facilitating fair, ethical, and reproducible research projects, there are also considerable challenges for the research team^10^. While research teams seek to deliver information in the informed consent process, there can be variability in information transmission from a research team’s representative in the consent conversation to the potential research participant, based on how the conversation unfolds^11^. Consent conversations are also very labor and time intensive, and limited to the working hours of study staff: this means scaling projects can be difficult in multi-site studies or for startup groups. Accordingly, this process has been identified as ripe for automation and innovation^10^.

Existing chatbots or “eConsent” systems are predominantly static portals that present PDF-like documents or linear multimedia^12,13^. While these approaches improve accessibility, they rarely provide reproducible conversational interaction or deterministic audit trails. Early pilots of chatbot-based consent in genomics demonstrated promise, but such tools remain exceptions rather than norms^14–16^.

For example, Schmidlen et al. 2019 stated that only ∼16% of participants had heard of chatbots and just ∼8% had ever used one, highlighting both the novelty of the approach and the need for further development. These pilots were developed within a commercial context (Invitae Corporation), and while they demonstrated feasibility, they were not designed for portability, reproducibility, or IRB audit across multiple institutions. Subsequent work in cascade testing^15^ and broader digital counseling^16^ reinforced that chatbot tools can improve engagement but are still limited in scope and rarely designed for reproducibility or cross-site auditability.

Regulatory guidance also stresses the importance of audit trails and version control in such eConsent systems^17^. Together, these observations underscore the gap: very few existing tools leverage conversational interfaces in a way that is both scalable and reproducible. Current options either lack scalability or lack the reproducibility and auditability that are essential in regulated biomedical research. In light of these challenges, we see an opportunity for a scalable, digital solution with the potential to improve the delivery of consent and inclusion in research - the chatbot, Kauro.

Kauro is a conversation graph-based chatbot that is fully auditable by an institutional review board (IRB) or ethics board that provides information transmission in place of a human representative from a research study team. Given the important humanistic aspect of the ritual of informed consent^18^, Kauro has key human backstops: 1) humans must design or approve the nodes of the conversation graph, 2) a potential research participant has, at each node, the option of an offramp to schedule a time to speak with a study staff member, and 3) if a quiz is not passed with a 100% score (10/10 questions right) a participant may not proceed with joining a research study without speaking with a study staff member. Such design considerations in a tool employs the clear utility of a chatbot without the risk of AI-LLM hallucination or missing information based on a variable human conversations (Table 1).

**Table 1:** Comparison of scripted and generative chatbots in the context of research consent. Scripted chatbots like Kauro rely on pre-approved JSON graphs for deterministic, auditable interactions that preserve IRB-approved language, while generative chatbots based on large language models (LLMs) offer greater flexibility but introduce variability, audit challenges, and compliance risks.

|  | Scripted Chatbot (Kauro) | Generative Chatbot (LLM-based) |
| --- | --- | --- |
| Source of content | Pre-approved IRB script, encoded as JSON graph | AI model trained on large text bodies |
| Behavior | Deterministic, node-by-node traversal | Stochastic, generates novel responses |
| Auditability | Fully auditable, reproducible, traceable | Difficult to audit; outputs vary with each run |
| Compliance | Ensures IRB-approved language is preserved | Risk of deviation from IRB content |
| Limitations | Cannot improvise; human staff needed for off-graph questions | Flexible but unpredictable; may hallucinate |

### Meet Kauro

Previous work, including ours, established a proof of concept for a scripted, quiz-gated, and IRB-approved chatbot^10,14^. That work demonstrated that conversational consent tools can be both feasible and effective. In our pilot^10^, which was deployed in support of the GREGoR Consortium^19^, participants using the chat tool completed sessions more quickly than in traditional staff-led workflows, achieved high scores on knowledge checks, and no adverse events were reported. This study, using a commercially available tool, established the viability of chat-based consent in genomic research.

Kauro builds on the foundation set in that pilot study, but with key innovations. Unlike currently available commercial tools, Kauro was designed in an academic setting as open-source software to ensure transparency, reproducibility, and portability across studies. Its modular architecture separates client, server, and graph (i.e., concerns), enabling adaptation to diverse research contexts. Conversation scripts are structured as JavaScript Object Notation (JSON)^20^ graphs with version control and audit trails, ensuring IRB-approved content is preserved across deployments. With extensible node types, automated testing, and built-in reproducibility, Kauro transforms the early promise of chat consent into a scalable, auditable, and generalizable platform for biomedical research.

### System Architecture

Kauro is a graph based chat engine where the content for a conversation is encoded in JavaScript Object notation (JSON) as a directed graph. Each conversation node in the graph has a definite structure (see Figure 1). This structure enables a deterministic traversal (the order of visitation is predictable and guaranteed to be the same every time for the same starting point) of the graph that is fully reproducible and auditable by investigators or institutional review boards. The transparency of the “conversation” with Kauro ensures adherence to IRB requirements.

**Figure 1:**
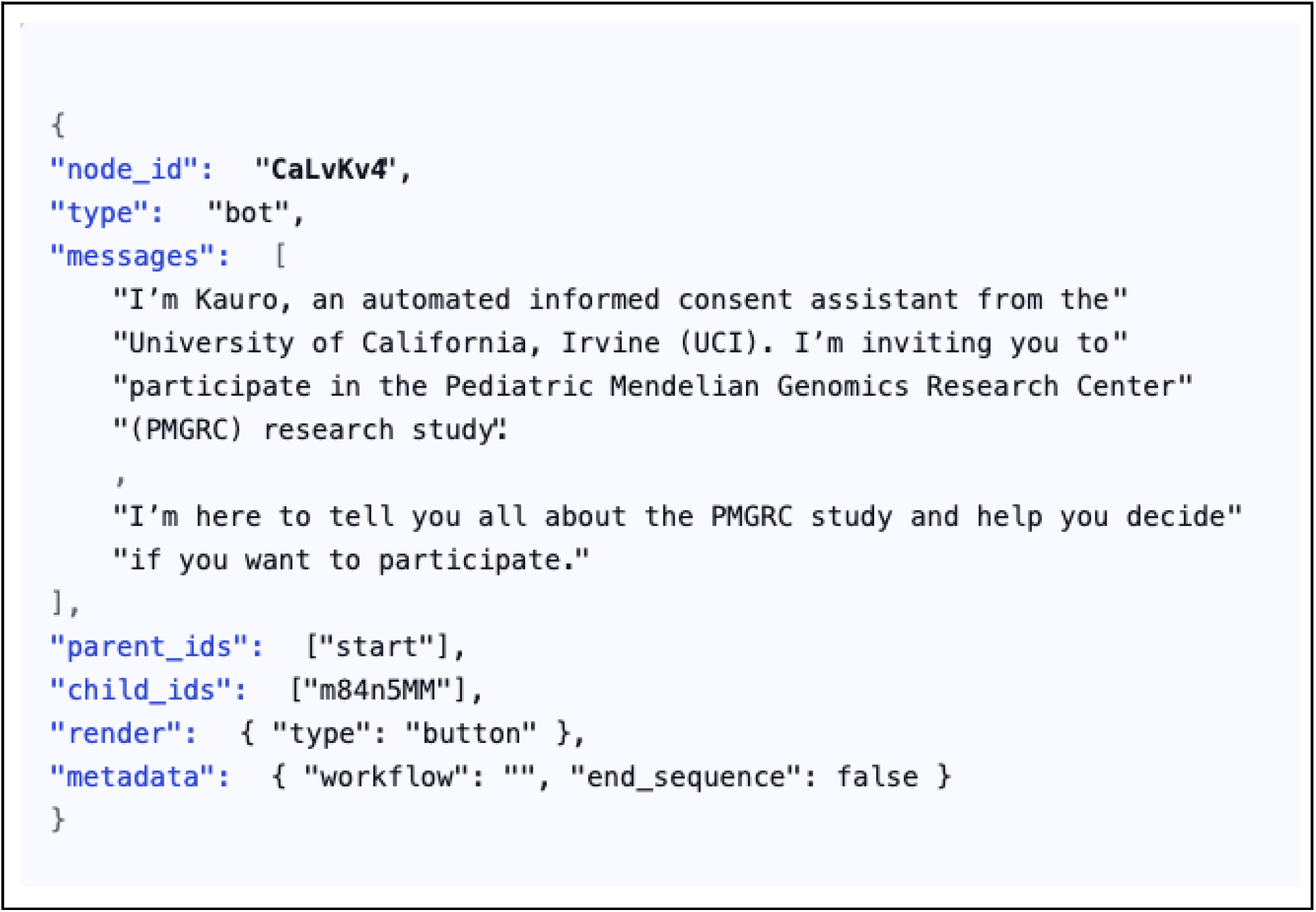
Stylized JSON representation of a Kauro node. Each node encodes its unique identifier, type (bot or user), parent and child links, and message payloads. Nodes may also contain rendering instructions (e.g., button, checkbox, text field) and metadata fields that guide workflow control, such as whether the node terminates a sequence. This schema enables deterministic traversal of the consent script, while semantic tags allow extended functionality such as scoring, alerts, or persistence. Together, these features make the node both human-readable and machine-executable, supporting reproducibility, version control, and auditability across deployments.

There are two critical innovations in Kauro: modularity and flexibility. The first innovation, modularity, is derived from the two main principles of *modular programming*^*21*^: “1) Allow one module to be written with little knowledge of the code in another, and 2) allow modules to be reassembled and replaced without reassembly of the whole system.” *Modular programming* is more often referred to as *Separation of Concerns (SoC)*^*22*^ in modern discussions, and Kauro accomplishes this by having the code for the client, the server, and the conversation graph as distinct modules, and able to be modified independently.

Second, as open source software, Kauro (released under the CC-BY-4.0 license) can be deployed in a variety of compute environments. Other off the shelf products (e.g., REDCap eConsent ^23^) require use of product sanctioned compute resources or are subscription services. Flexibility in deployment has notable advantages for control of the data and auditability by a given institution or firm that chooses to deploy Kauro.

In terms of SoC, there are three concerns for Kauro: the client, the server, and the conversation graph. The client, or user interface, provides a way for the user to interact with the content from the server. The server, or backend, processes the user responses and then returns a new node to the user. This strict representational state transfer (REST) application programming interface (API) contract design contrasts with the many public REST APIs, which frequently diverge from REST guidelines and lack consistent error handling or version control^24,25^. By enforcing schema-driven inputs and deterministic outputs, Kauro ensures auditability and reproducibility even as other API ecosystems struggle with consistency.

Such functionality is not limited to interactions that originate with Kauro users via the client. The study staff who create the chat graph, manage participant enrollment, and perform subsequent data analysis are all subject to this aspect of Kauro SoC functionality. For emphasis, any properly-formatted request (i.e. an API call as defined by the documentation (https://w3id.org/UCI-ICTS/kauro)) can be processed by the server with a return response. This allows any server instance to have interactions with other applications - for example, RedCAP.

Each concern is independent of the others and can be adjusted without disrupting functionality. The system can be deployed in a variety of environments, and the source code and instructions are available on GitHub at (https://w3id.org/UCI-ICTS/kauro). The conversation graphs are portable JSON objects that can be version controlled, shared, modified, and audited. Kauro can function with any number of graphs, as long as they conform to the predefined structure (Figure 1). The combination of the Kauro predefined structure and the generalizability of JSON objects enables as many different projects as a deployment site has project conversation graphs.

The node schema (Figure 1) is minimally constrained to allow extensibility. Beyond simple question & answer interactions, nodes may define forms (radio buttons, checkboxes, free text, dropdowns). Semantic tags further extend functionality by linking nodes to services such as scoring, alerts, and data persistence. This design allows research teams to adapt Kauro to diverse protocols without modifying the underlying engine.

The backend, implemented in Django^26^, serves as the execution engine. Its function is to enforce the graph: user requests are mapped to node identifiers, validated against schema, and deterministically resolved to the next node. This strict enforcement ensures that content remains IRB-approved, while schema-driven validation guarantees reproducibility across deployments.

The client, implemented in React ^27^, dynamically renders nodes returned by the server. It accommodates varied content types (forms, inputs, informational messages) and presents them responsively across devices. Because the client is decoupled from the graph content, any properly formatted script can be rendered without modification to the interface.

As outlined in Figure 2, these design choices yield core features critical for consent workflows: strict version control; deterministic graph traversal; dynamic rendering of varied input types; session persistence to allow participants to pause and resume; full audit logging of all interactions; and support for multiple user roles (participants, guardians, administrators, staff)

**Figure 2.**
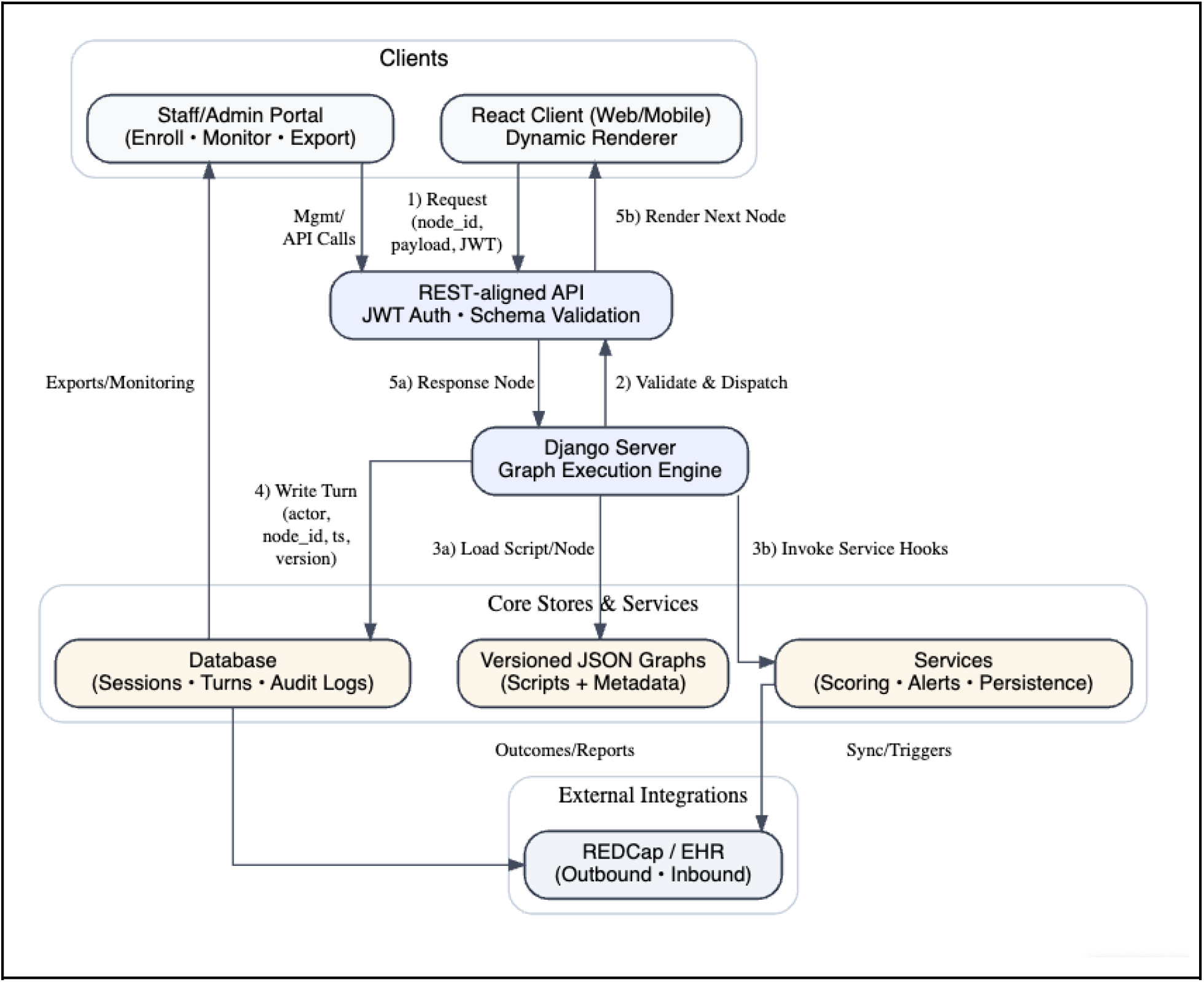
Data flow diagram of the Kauro system. Participants interact through a React client, while study staff use an administrative portal for enrollment and monitoring. All interactions pass through a REST-aligned API that enforces authentication and schema validation before reaching the Django execution engine, which interprets the IRB-approved conversation graph. The engine retrieves versioned JSON scripts, invokes modular services such as scoring and alerts, and records each turn to a database of sessions and audit logs. Responses are returned deterministically to the client for rendering. Staff may export data directly, and selected outcomes can be integrated with external systems such as REDCap or electronic health records. Numbered arrows indicate the request–response cycle: (1) client request; (2) API validation and dispatch; (3a) script retrieval and (3b) service invocation; (4) audit logging; and (5a) response and (5b) rendering.

### Reproducibility

Each consent chat graph has to be authored by domain experts. In our experience for informed consent, this can be achieved by a collaborative team of genetic counselors, physicians, and research scientists. Different conversations will require different human design teams. Before deployment each chat graph undergoes formal review and approval by an Institutional Review Board (IRB), just like a traditional paper consent form or other multimedia consent modules.

Kauro versions each JSON graph with the graph name (filename) and the IRB approval reference. This ensures that participants are always presented with the exact IRB-approved chat^17^. Every chat session record is linked to the originating graph and IRB reference ID, creating a deterministic audit trail. Investigators can reconstruct the precise route traversed, seeing what the participant saw, what responses were given, and which version of the consent protocol was assigned to that participant. Such strict versioning contrasts with most commercial or ad hoc digital consent tools, which often treat content updates as opaque and lack formal auditability. In multi-site studies, where reproducibility across deployments is essential, version control provides a shared reference point that enables investigators and regulators to confirm consistency of the consent experience across institutions and time^28,29^.

The core engine of the chatbot Kauro is graph-based, not probabilistic. The predetermined graph traversal rules that run the engine serve as a guarantee that the same input leads to the same output. The absence of adaptive learning prevents any deviation from the specific IRB approved output. Such a deterministic traversal, in contrast to a probabilistic or adaptive system, protects the IRB-approved content from drift or evolution during deployment. Each session is explicitly linked to a graph version (graph name and the IRB approval reference), as described above.

Every chat is logged with a set of standardized fields: actor (bot or user), node_id, payload, timestamp, and graph_version. This schema creates a complete, immutable audit trail. Such audit trails are not only a best practice in software engineering but also a regulatory requirement: FDA Part 11 guidance^17^ and EMA expectations^30^ specify that electronic records must preserve unalterable audit logs.

Empirical reviews of eConsent platforms have shown many current systems provide only partial logging, if any^12,13,23^. Kauro explicitly addresses this gap by coupling every chat turn with script version and IRB reference, enabling reproducibility across sites and across longitudinal studies. This structured logging allows research teams to verify comprehension scores, document who consented to what, and track escalations or follow-ups. The administration dashboard leverages this information to provide cross-study oversight, audit preparation, and regulatory reporting, turning technical logs into operational tools for compliance.

### Validation

In addition to integration and regression tests, Kauro employs unit tests to validate individual node structures, schema conformance, and service hooks. All test suites run automatically in a continuous integration pipeline^31^, ensuring that any change to the client, server, or conversation graphs does not alter approved traversal behavior. Together, these measures make validation reproducible across deployments and across software updates^32,33^.

Technical reproducibility alone is not sufficient in regulated biomedical research; robust governance ensures that consent processes remain trustworthy across sites and over time. Kauro incorporates governance at several levels. At the content level, only designated study staff with appropriate permissions may author or edit conversation graphs. Proposed revisions are version-controlled and subject to IRB approval before deployment, ensuring that no content reaches participants prior to approval.

At the operational level, administrative or user role-based access control distinguishes between participants^34^, guardians, study staff, and administrators. Each role has restricted privileges: participants can only interact with their assigned session; staff can monitor progress and export audit logs; administrators can manage deployments. These controls protect against accidental or unauthorized modifications.

At the oversight level, every chat is recorded with graph version and IRB reference, enabling independent review by regulators^17,30^, data safety monitoring boards, or cross-site coordinating centers. The administrative dashboard aggregates this information to provide a transparent view of consent activity in real time.

Finally, Kauro’s open-source release under a permissive license allows external stakeholders to inspect, validate, and extend the system^35,36^. This combination of access control, audit trails, and transparency links technical validation with organizational accountability, meeting the demands of reproducibility and compliance.

### Implementation for rare disease participant recruitment

The first use case for Kauro is in support of the University of California, Irvine GREGoR site. GREGoR is a large-scale rare and Mendelian condition consortium that seeks to systematize the discovery and diagnosis of such conditions. Importantly, GREGoR focuses on exome and diagnosis-negative cases. Given the gap between the high number of referrals to the consortium and the targeted nature of recruitment for conditions with a likelihood of genetic etiology, this was an ideal case for the implementation of this chatbot.

The script was based off of previously published literature and modified to our site’s specific details ^10^. Important system guardrails implemented in this iteration of Kauro were a 10-question quiz with consent only allowed after 100% of questions were answered correctly. In the case of missed questions, a teach-back system was implemented where another chance would be given to answer questions after right answers were provided. The chat script allows for users of the age of majority to fill out consent for themselves and their legal guardians.

This script is maintained in JSON format by Kauro (Supplement 1). Kauro has additional functionality to export pdfs for review (Supplement 2). While JSON can be interpreted by human readers, these pdfs provide a much easier document for review by IRBs and human users, while preserving the content presented in the chats and the informatic backbone - specifically node and edge IDs (in parent and child relationship notation). The site IRB approved this process in totality with the supplemental documents under IRB #4469 - Pediatric Mendelian Genomics Research Center.

## Discussion

### Why structured/non-learning AI matters

Kauro, a pre-defined graph-based conversational chatbot, presents an exciting, ethically-grounded alternative to adaptive, probabilistic, LLMs for information transmission to research participants in the biomedical enterprise. Prior work in software engineering similarly emphasized that deterministic chatbots, while perhaps less engaging, offer predictability and safety^37^. Kauro leverages this principle in a biomedical context, where reproducibility and auditability outweigh conversational novelty. While we initially designed Kauro for informed consent, it is generalizable to any conversation where predetermined, standardized information must be conveyed with a high degree of accuracy.

Early chatbot pilots in genomics, such as those developed by Invitae Corporation^10,14,15^, demonstrated the feasibility and participant acceptance of scripted, graph-based consent. Such implementations were proprietary and not designed for portability, reproducibility, or multi-institutional audit. Kauro was developed in an academic setting, released under an open-source license, and designed with modularity and version control at its core. This shift from commercial proof-of-concept to academic infrastructure highlights how the same conceptual model, a scripted chatbot, can evolve into a transparent, auditable, and generalizable platform suitable for biomedical research and medicine.

LLMs are trained on large collections and are able to interpret natural language input from users. This can offer great flexibility in their ability to respond to users. It also enables natural language input. However these benefits come with tradeoffs. The LLM chatbots by definition are not predictable; they are probabilistic. In situations where consistency and clarity are favored, or required like in informed consent, this is not ideal^17,30^.

Kauro is a deterministic, graph-based tool. Such determinism ensures no hallucinations or deviation. The absence of a backend powered by a probabilistic model is, perhaps counterintuitively, the innovation. Every response (from Kauro, or from the user) is pre-vetted. This ensures reproducibility, auditability, and compliance. Kauro shows that the core features desired by the use of machine intelligence (automation, structured reasoning, branching logic) can be used without having to use probabilistic models.

### Safety and constraint by design

Kauro’s constraint-by-design reflects a principle long recognized in safety-critical systems: reliability often depends on limiting flexibility. Kauro can serve many participants in parallel. The chat graphs are portable across studies and sites. While an LLM may offer a dynamic experience and be able to interact with a broader range of questions, this introduces the risk of straying from approved content, which may be harmful to the participant and hinder information transmission. Kauro’s constraint by design does not allow for unexpected user questions, and the resultant possibility of unexpected bot responses. This is acceptable because the user always has the option to reach out to the study staff.

Kauro provides a case study for rigorous application programming (API) engineering in biomedicine. Analyses of public Representational State Transfer (REST) APIs reveal that fewer than 1% fully comply with REST principles and that error handling, versioning, and documentation remain problematic across the ecosystem^24,25^. While Kauro does not implement every aspect of REST (e.g., Hypermedia as the Engine of Application State/HATEOAS), its API emphasizes schema validation, deterministic outputs, and explicit versioning - practices that empirical studies show are underutilized in most public APIs. These principles may serve as a model for other safety-critical applications where reproducibility and auditability are paramount. By embedding schema validation and versioning directly into the API layer, Kauro demonstrates how biomedical software can avoid the pitfalls identified in broader surveys of REST practice^13,14^.

Implementation of a modified peer-reviewed consent script and subsequent IRB approval emphasizes both the generalizability of this approach and the suitability of this method for compliance with US laws and regulations for human subjects protection. We are confident that this is the first step to an internationally-agnostic tool for particular regulated interactions between researchers and potential participants.

### Challenges

Several challenges remain. First is user acceptance. Some participants may conflate any chatbot with artificial intelligence more broadly and be skeptical of delegating a critical process such as consent to what they perceive as “AI”. This skepticism is amplified by well-publicized failures of generative models^38^, even though Kauro’s design is fundamentally different. Clear framing that emphasizes Kauro’s deterministic nature, coupled with the availability of study staff for follow-up, will be essential to maintaining trust^10,14,15^.

Another challenge is rooted in access^39^. Kauro was designed for mobile-first interaction, making it flexible and convenient for many participants. However, the platform depends on internet connectivity, and disparities in access to reliable broadband or devices could exclude certain potential users. These issues are not unique to Kauro but reflect a broader challenge in digital health implementation: ensuring that scalable tools do not inadvertently reinforce existing inequities^40^.

Finally, maintenance overhead presents a barrier. Each revision of a conversation graph should require renewed IRB approval, a process that is necessarily rigorous but time consuming. This constraint can slow the pace of iteration compared to commercial digital platforms. Yet, paradoxically, it is also what guarantees Kauro’s reproducibility and auditability. Far from being a weakness, this cycle of approval and version control underscores Kauro’s commitment to regulatory compliance.

### Future directions

Despite the challenges outlined above, there are different paths forward for extending Kauro’s impact that are reassuring. Hybrid approaches that combine the deterministic backbone with tightly constrained generative models may allow for a more natural feeling question-and-answer. Such guardrails could allow participants to ask clarifying questions in their own words, while ensuring that answers are drawn from pre-approved sources and never modify the core consent script^38,41^. A chatbot that can readily and generatively say “I don’t know, but I can connect you to someone to help” would be an innovation to assuage key concerns.

Another critical direction is the development of multilingual scripts. By authoring and validating consent graphs in multiple languages, Kauro can broaden participation and improve global research participation^40^. This aligns with broader initiatives to reduce barriers to research representing broader populations^17^.

Integration into clinical workflows also represents a natural next step. Linking consent outcomes directly to electronic health records or biobank pipelines would make Kauro not only a tool for recruitment but also an operational bridge between research and care delivery. Such integration would reduce duplication of effort and provide a richer data environment for investigators and clinicians alike.

Finally, Kauro’s underlying framework is generalizable. The same principles of deterministic traversal, auditability, and modular extensibility can apply to informational transmission in clinical trials, disease registries, and public health initiatives. By providing a transparent and reproducible conversation infrastructure, Kauro offers a template for digital conversations across biomedical domains where safety, compliance, and trust are paramount^12,13^.

## Conclusion

Informed consent remains a high-stakes process: complex content, variable delivery, and substantial operational burden across sites. While generative AI has energized conversational interfaces, its probabilistic outputs and potential to deviate from approved language make it ill-suited to IRB-regulated consent conversations, or other such conversations where high fidelity transmission of information is paramount.

The graph-based chatbot Kauro offers a different path. By encoding a conversation as a versioned JSON graph and enforcing deterministic traversal, Kauro preserves IRB-approved content verbatim while enabling scalable, script-driven conversations. The separation of concerns between client, server, and script, combined with an open-source codebase supports portability across institutions and transparent review by investigators and auditors.Karuo thus reframes “intelligence” for consent as precision and proof: deterministic delivery, complete audit trails, and verifiable compliance at scale.

## Data Availability

This study was funded by GREGoR U01HG011745 and ICTS UM1TR004927

https://github.com/UCI-ICTS/kauro

