## Supplemental file 2 for "Kauro, a graph-based chatbot for high-fidelity information transmission conversations"

### Full Consent Chat Transcript

#### Main

**Node ID:** CaLvKv4

**Parents:** start

**Children:** m84n5MM

**Kauro:** I'm Kauro, your automated Medical Information Assistant from the University of California, Irvine (UCI). I'm inviting you to participate in the **Pediatric Mendelian Genomics Research Center (PMGRC)** research study.

**Kauro:** I'm here to tell you all about the PMGRC study and help you decide if you want to participate

---

**Node ID:** m84n5MM

**Parents:** CaLvKv4

**Children:** de5ZDgm

**User Transcript:** 👋 Hi Kauro!

---

**Node ID:** de5ZDgm

**Parents:** m84n5MM

**Children:** PJwkbgi

**Kauro:** Some of the things I will review with you are: why we are doing this research, what you will be expected to do if you enroll, and possible risks and benefits

**Kauro:** This information will help you choose whether or not to participate

---

**Node ID:** PJwkbgi

**Parents:** de5ZDgm

**Children:** oS9m8Ua

**User Transcript:** Great!

---

**Node ID:** oS9m8Ua

**Parents:** PJwkbgi

**Children:** me5RCBw, iLpXdAS

**Kauro:** Our conversation usually takes about 30 minutes. At the end, if you choose to enroll, you can sign the consent.

**Kauro:** This link will expire two weeks after it was sent to you. You can always take a break and come back to this link, during the two weeks, to complete enrollment.

---

**Node ID:** me5RCBw

**Parents:** oS9m8Ua

**Children:** a6JKRdh

**User Transcript:** Okay

---

**Node ID:** a6JKRdh

**Parents:** iLpXdAS, me5RCBw

**Children:** WwRBQik

**Kauro:** All of the information we discuss will also be described in more detail in our consent document

**Kauro:** You can only sign the consent form electronically after reviewing it with me or with a study team member

**Kauro:** You can access an example of the consent document by clicking this link, but please remember to return here to complete our chat

---

**Node ID:** WwRBQik

**Parents:** a6JKRdh

**Children:** XcPGk57

**User Transcript:** Can I look at the consent document after we chat?

---

**Node ID:** XcPGk57

**Parents:** WwRBQik

**Children:** JPVkFCr

**Kauro:** Yes, you can come back here to access the link later, and I can also provide the link again at the end of the chat

---

**Node ID:** JPVkFCr

**Parents:** XcPGk57

**Children:** ZjTLunQ

**User Transcript:** Okay, great

---

**Node ID:** ZjTLunQ

**Parents:** JPVkFCr

**Children:** fWXv5QS

**Kauro:** Whether or not you choose to participate in the study, the team at UCI may review your interactions with me, Kauro, to better understand how people interact with me and how I can improve

---

**Node ID:** fWXv5QS

**Parents:** ZjTLunQ

**Children:** S2nH9zZ

**User Transcript:** Ok, sounds good

---

**Node ID:** S2nH9zZ

**Parents:** fWXv5QS

**Children:** 9DgwYTz, DdxFUae, BxD4wrq

**Kauro:** If you are ready, we can start going through the information together right now!

**Kauro:** If you would rather review the consent later or you would rather review it with a study team member, just let me know!

---

**Node ID:** 9DgwYTz

**Parents:** S2nH9zZ

**Children:** Q8yaxQc

**User Transcript:** I'd like to continue chatting with Kauro now

---

**Node ID:** Q8yaxQc

**Parents:** 9DgwYTz, 87q8gWG

**Children:** M4CaAt5

**Kauro:** Okay!

**Kauro:** Let's do this!

**Kauro:** Please be sure to take notes and write down any questions you have as we go through this chat. You will have the option to talk to a study team member at the end. ✏️

**Kauro:** If you have any questions that are not answered during this chat, we will put you in touch with a team member to discuss them further

---

**Node ID:** M4CaAt5

**Parents:** Q8yaxQc

**Children:** FADCStr

**User Transcript:** Sounds good! 👍

---

**Node ID:** FADCStr

**Parents:** M4CaAt5

**Children:** Vew9XnD, Bkyp7hD

**Kauro:** The goal of this study is to help identify the cause or causes of genetic health conditions

**Kauro:** You are being invited to join this study because you or someone in your family has a suspected genetic health condition (or a health issue that might be related to genes)

**Kauro:** We often enroll multiple family members (people related by blood) in the study

---

**Node ID:** Vew9XnD

**Parents:** FADCStr

**Children:** KMirb2i

**User Transcript:** Okay

---

**Node ID:** KMirb2i

**Parents:** RXBDyqd, Vew9XnD

**Children:** dM9iaFR

**Kauro:** Ideally, we would enroll **the person with the suspected genetic health condition** as well as **both of their biological parents** (mom and dad), if possible.

**Kauro:** So, if one or both parents are able to enroll, that would be great!

**Kauro:** You can still participate if family members are not available

---

**Node ID:** dM9iaFR

**Parents:** KMirb2i

**Children:** E3C8G5Y

**User Transcript:** Good to know!

---

**Node ID:** E3C8G5Y

**Parents:** dM9iaFR

**Children:** C8baGp6

**Kauro:** Each person who enrolls in the study will have to go through this study consent process

**Kauro:** You can enroll yourself and/or your children in this chat today. We can send a separate chat link to any other adults who want to enroll

---

**Node ID:** C8baGp6

**Parents:** E3C8G5Y

**Children:** CPf9CCz

**User Transcript:** Okay

---

**Node ID:** CPf9CCz

**Parents:** C8baGp6

**Children:** BBfTzRw, CAwKwCW

**Kauro:** For children who are 6 years old or younger, a parent or guardian can complete the consent on their behalf

**Kauro:** For children ages 7 to 17, (or for adults who are under a person's legal guardianship), a parent or guardian must complete the official consent

**Kauro:** We will also need to talk directly to the child (if they are capable) to get their agreement about participating. This is called assent

**Kauro:** People who are age 18 or older need to go through the consent process themselves unless they are under another person's legal guardianship

---

**Node ID:** BBfTzRw

**Parents:** CPf9CCz

**Children:** USfk36C

**User Transcript:** Okay

---

**Node ID:** USfk36C

**Parents:** CAwKwCW, BBfTzRw

**Children:** b5nYNf6

**Kauro:** Can you tell me who in your family might consider enrolling in this study? (check all that apply, including any parents or other adults who might enroll)

---

**Node ID:** Ey8gJrc

**Parents:** L5xPJtN

**Children:** KR89wri, QXNa2cz

**Kauro:** You're welcome 😀

**Kauro:** Now, let's take a moment to talk a little bit about genetics.

**Kauro:** Do you want to keep it simple or get a little more science-y?

---

**Node ID:** KR89wri

**Parents:** Ey8gJrc, WnXoffX

**Children:** hBC6sGH

**User Transcript:** Science-y please 🤓

---

**Node ID:** hBC6sGH

**Parents:** KR89wri

**Children:** EcgsbMN

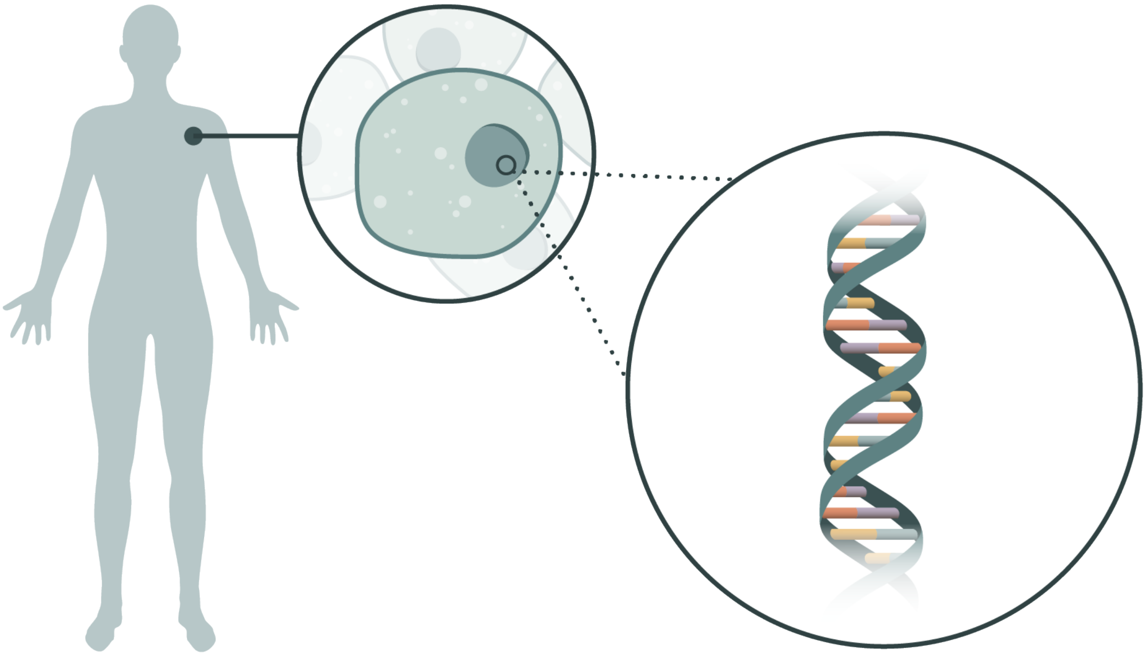

**Kauro:** I was hoping you would say that!

**Kauro:** You may already know that your body is made of trillions of cells. DNA provides the instructions to tell our bodies how to grow and develop

---

**Node ID:** EcgsbMN

**Parents:** hBC6sGH

**Children:** SfPWkep

**User Transcript:** Ah, okay

---

**Node ID:** SfPWkep

**Parents:** EcgsbMN

**Children:** AonpoZW

**Kauro:** Our DNA uses specific letters (A, T, C, and G) to write these instructions

**Kauro:** A little section of the code might look something like this:

ATGCGCTAGCTCCTAGCTAGCCTAGCTAGCTAACGCTAGCCTCGTAGACTACGATCGCTAAGCTAGC

**Kauro:** Our genetic code is actually over 3 billion letters long!

---

**Node ID:** AonpoZW

**Parents:** SfPWkep

**Children:** 45kz8CE

**User Transcript:** That's complicated

---

**Node ID:** 45kz8CE

**Parents:** AonpoZW

**Children:** QjTtL5s, 9ZmfvBQ

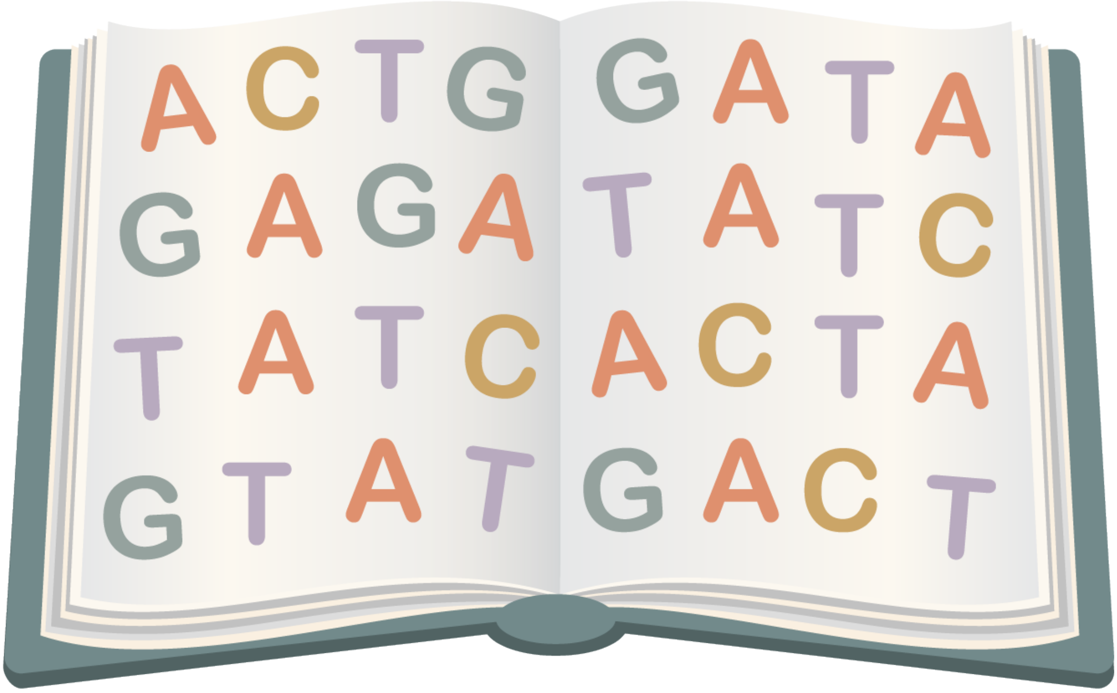


**Kauro:** I know this sounds complicated! Your cells know how the read these instructions. They use your genetic code like a cookbook

**Kauro:** Our DNA is organized into specific sections, as if they are individual recipes. These sections, or the recipes, are called genes

---

**Node ID:** QjTtL5s

**Parents:** 45kz8CE

**Children:** BF3k6W7

**User Transcript:** This sounds familiar

---

**Node ID:** BF3k6W7

**Parents:** 9ZmfvBQ, QjTtL5s

**Children:** 8Pvoowc

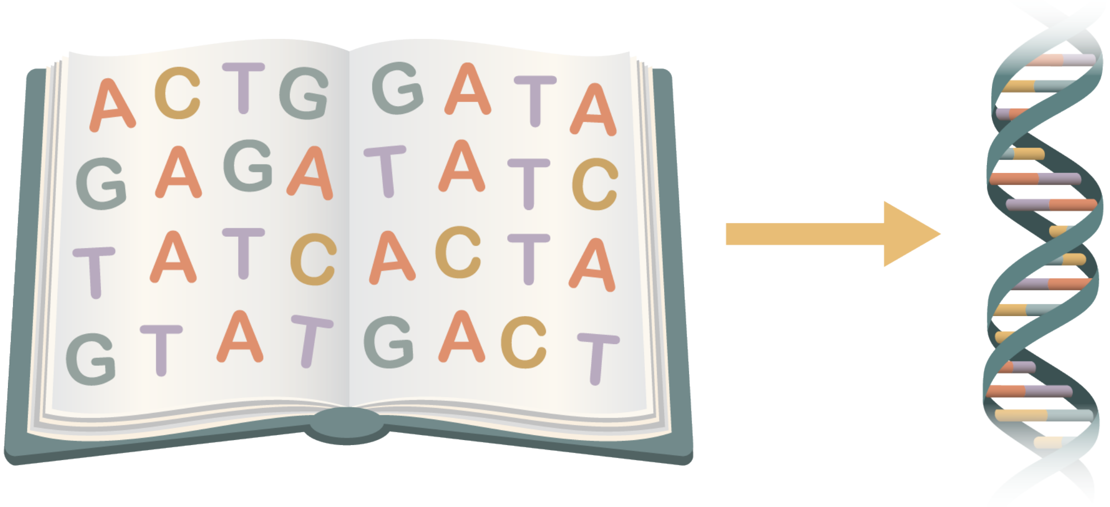


**Kauro:** In a cookbook, each recipe gives instructions for making a different dish

**Kauro:** In your cells, each gene provides the instructions for molecules called RNA, and each RNA makes a different protein

---

**Node ID:** 8Pvoowc

**Parents:** BF3k6W7

**Children:** WQFz456

**User Transcript:** That’s interesting

---

**Node ID:** WQFz456

**Parents:** 8Pvoowc

**Children:** kvTbHhw

**Kauro:** Your body uses proteins to do many important things

**Kauro:** For example, some proteins build our tissues, like muscles and nerves. Other proteins help our body break down food and make energy

---

**Node ID:** kvTbHhw

**Parents:** WQFz456

**Children:** PpUbXu4

**User Transcript:** That’s cool!

---

**Node ID:** PpUbXu4

**Parents:** kvTbHhw

**Children:** TfVxQFn

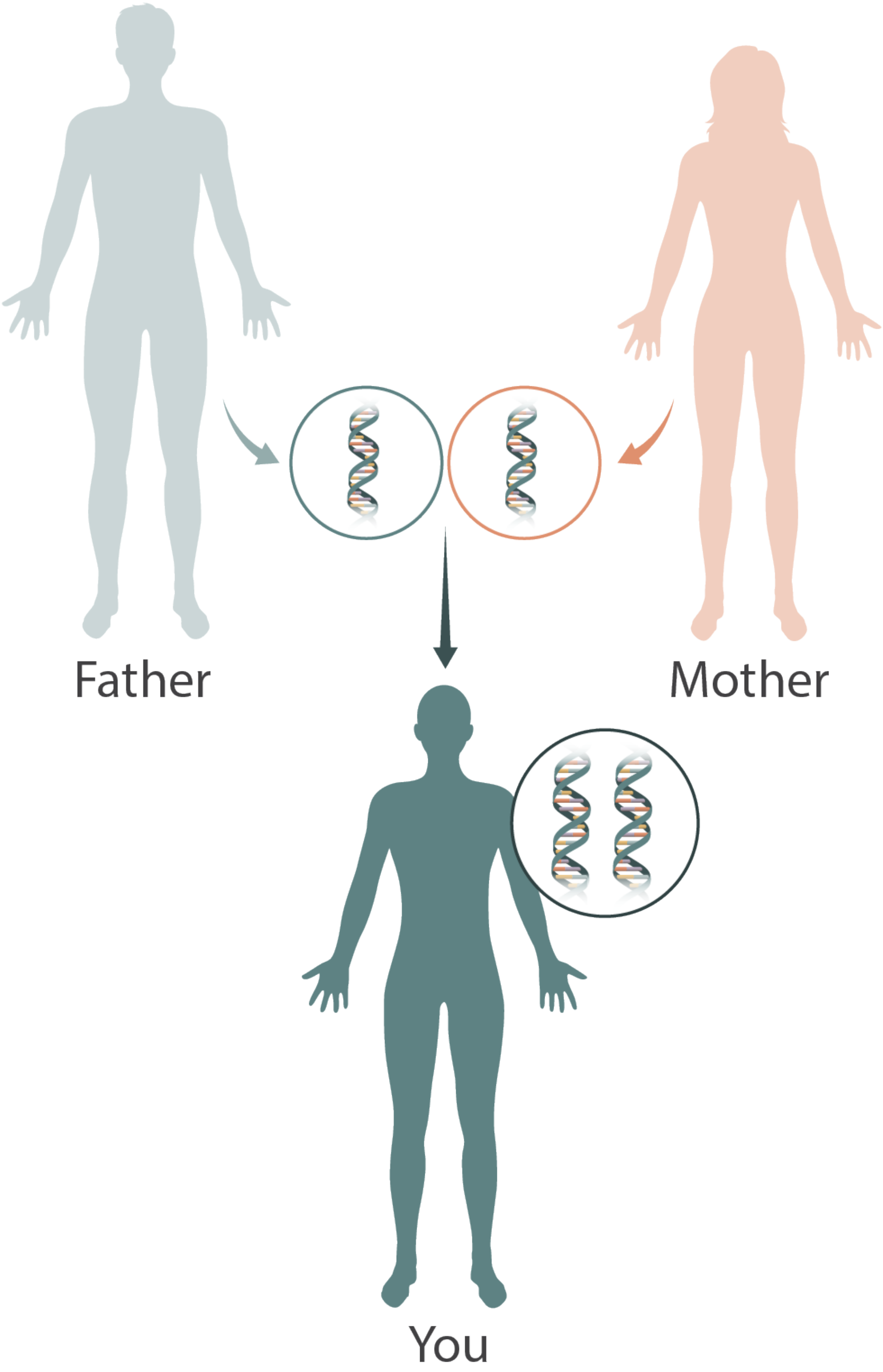


**Kauro:** I know, right?

**Kauro:** You typically have 2 copies of each gene - getting one from each of your parents. The 2 copies work together to make you who you are

---

**Node ID:** TfVxQFn

**Parents:** PpUbXu4

**Children:** HVeQxH2

**User Transcript:** Got it

---

**Node ID:** HVeQxH2

**Parents:** TfVxQFn

**Children:** Vnd4AC6

**Kauro:** People can have different versions of the same gene. This is what makes each person unique

**Kauro:** We call these different versions of a gene “variants”

---

**Node ID:** Vnd4AC6

**Parents:** HVeQxH2

**Children:** c85Yw2k

**User Transcript:** What is a variant?

---

**Node ID:** c85Yw2k

**Parents:** Vnd4AC6

**Children:** iumYvk4

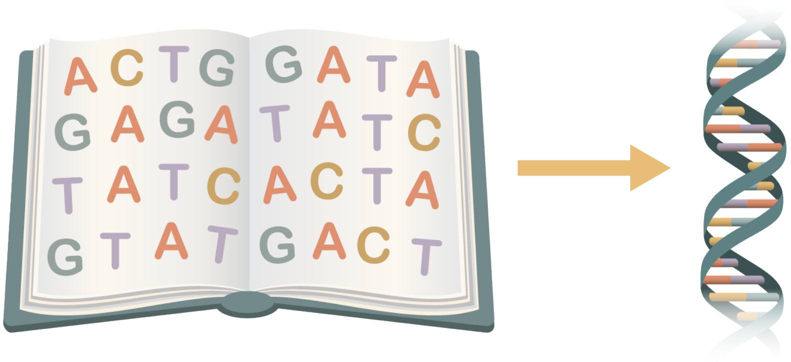

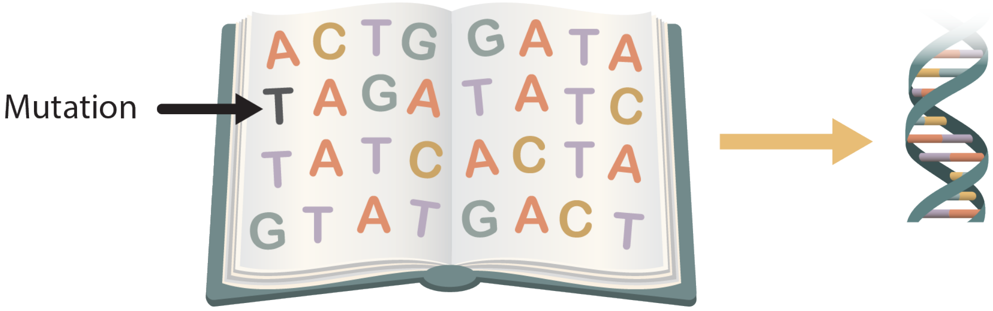


**Kauro:** Remember how we said your genes are like recipes?

**Kauro:** When genes have misspellings or changes in them, we call that a variant. This is like changing an ingredient in a recipe. The end product may turn out differently than you expect or may not work at all

---

**Node ID:** iumYvk4

**Parents:** c85Yw2k

**Children:** 7i6JiiZ

**User Transcript:** I see

---

**Node ID:** 7i6JiiZ

**Parents:** iumYvk4

**Children:** ZcynmTH

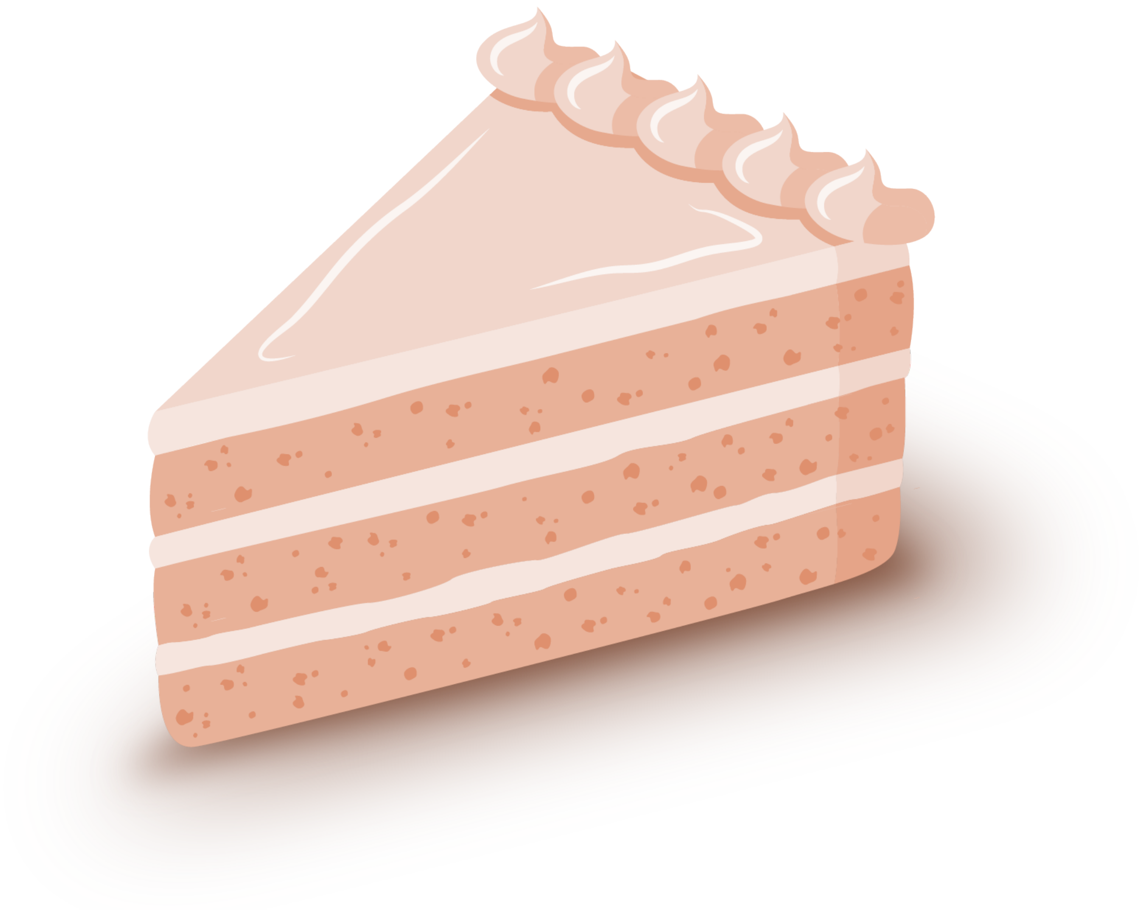

**Kauro:** Imagine that you are baking a cake and the recipe tells you to use salt instead of sugar, or strawberries instead of chocolate!

**Kauro:** That's going to affect the cake!

---

**Node ID:** ZcynmTH

**Parents:** 7i6JiiZ

**Children:** EE6kxBa

**User Transcript:** Got it

---

**Node ID:** EE6kxBa

**Parents:** Ti3ZavP, ZcynmTH

**Children:** iMcrZ9h, iVCEBuL

**Kauro:** Genetic variants are what make each person unique

**Kauro:** Some variants may change a person's eye color, height, or hair color, while others may change a person's development, heart function, or kidney function

---

**Node ID:** iMcrZ9h

**Parents:** EE6kxBa

**Children:** 8Z6qtgu

**User Transcript:** Interesting

---

**Node ID:** 8Z6qtgu

**Parents:** iVCEBuL, iMcrZ9h

**Children:** Zgg7vB9

**Kauro:** When someone has a health issue or symptoms caused by a genetic change (or “variant”), this can cause a genetic disorder

**Kauro:** When we suspect that a patient has a genetic disorder, we often do genetic testing to try and find the change, or variant, that is causing the symptoms

**Kauro:** In the PMGRC study, we are interested in doing different kinds of genetic testing, some that is not available to your doctors regularly, to look for changes that might explain your health concerns

---

**Node ID:** Zgg7vB9

**Parents:** 8Z6qtgu

**Children:** iCSWyTS

**User Transcript:** Great

---

**Node ID:** iCSWyTS

**Parents:** Zgg7vB9

**Children:** GLzHadvB

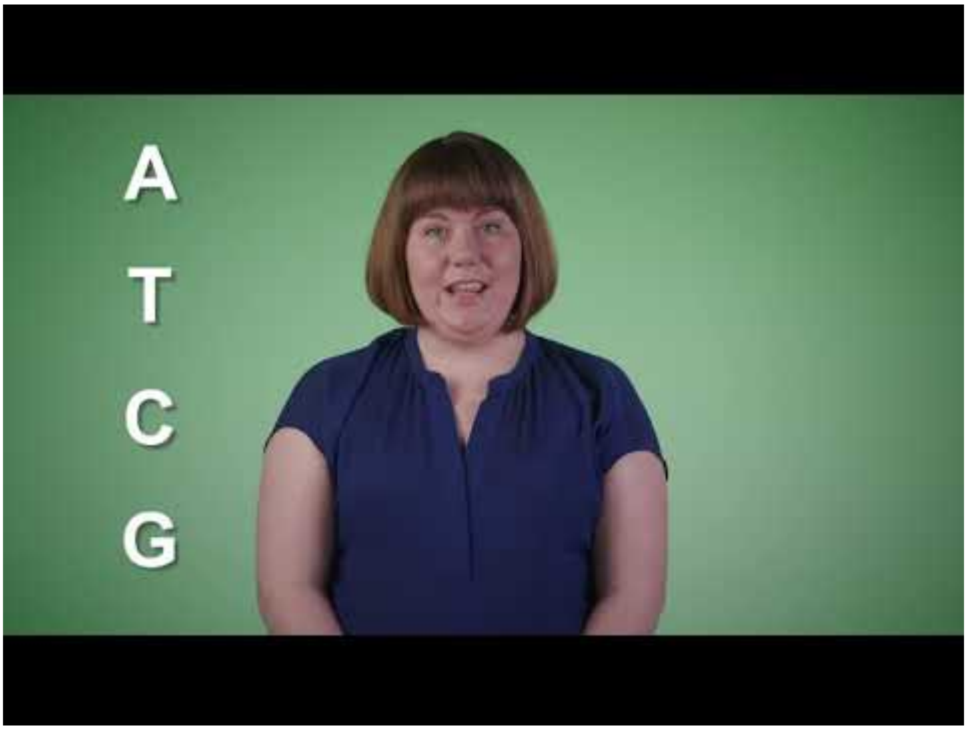


**Kauro:** Video link: https://www.youtube-nocookie.com/embed/jhJW69Ols14

**Kauro:** Here's a 1-minute video that talks about one common type of testing, called sequencing

---

**Node ID:** GLzHadvB

**Parents:** iCSWyTS

**Children:** jczufDn

**User Transcript:** Interesting

---

**Node ID:** jczufDn

**Parents:** iCSWyTS, GLzHadvB

**Children:** GLzDUvB, 9cuqxwT

**Kauro:** As we discussed earlier, we are hoping to enroll patients with known or suspected genetic conditions and their blood relatives, such as parents and siblings

**Kauro:** Can you tell me a little bit more about your history with genetic testing for you or your family members so far?

---

**Node ID:** GLzDUvB

**Parents:** jczufDn

**Children:** G5dyfh6

**User Transcript:** Never had genetic testing / I don't recall...

---

**Node ID:** G5dyfh6

**Parents:** GLzDUvB

**Children:** FiRFHGF

**Kauro:** No problem!

**Kauro:** One goal for our PMGRC study is to discover **new** causes of health problems, like finding a new gene or new ways that differences in our genes can cause health concerns

**Kauro:** If genetic testing didn't find an answer before, our research might be able to find something new

---

**Node ID:** FiRFHGF

**Parents:** kYjpLjd, G5dyfh6

**Children:** SgDDtr8

**User Transcript:** Okay...

---

**Node ID:** SgDDtr8

**Parents:** FiRFHGF

**Children:** hxyY83b, LbgZ4YK

**Kauro:** Another goal of our study is to better understand prior genetic test results that may have been unclear

**Kauro:** For example, if previous genetic testing found a “variant of uncertain significance”, our research might be able to help explain whether that finding is related to your symptoms or not

---

**Node ID:** hxyY83b

**Parents:** SgDDtr8

**Children:** XKshQvX

**User Transcript:** Got it!

---

**Node ID:** XKshQvX

**Parents:** DKTqMXc, hxyY83b

**Children:** oNfduXy

**Kauro:** We don’t know if this kind of research genetic testing will help people find a diagnosis, but we hope that it will at least help make science and medicine better with the possibility of helping other people in the future

---

**Node ID:** oNfduXy

**Parents:** XKshQvX

**Children:** GokRRmf

**User Transcript:** Well that sounds nice

---

**Node ID:** GokRRmf

**Parents:** oNfduXy

**Children:** HEz7sik

**Kauro:** We want to make sure you know that joining this PMGRC study is **voluntary**

---

**Node ID:** HEz7sik

**Parents:** GokRRmf

**Children:** NHPvQRe

**User Transcript:** What does that mean?

---

**Node ID:** NHPvQRe

**Parents:** HEz7sik

**Children:** Np4Ajah, HUAk3zG, Ua23Tdf

**Kauro:** Voluntary means you can choose to be part of this study or choose not to be part of this study. If you choose to enroll, then later change your mind, you can withdraw from the study

**Kauro:** Whatever choice you make, the quality of care you receive from your current medical team will not change. You will not lose access to your medical care or give up any of your legal rights or medical benefits

**Kauro:** If we make any changes to the study or if there are any important new findings, we will tell you so that you can decide if you still want to participate

---

**Node ID:** Np4Ajah

**Parents:** NHPvQRe

**Children:** ckDLoqi

**User Transcript:** Sounds good!

---

**Node ID:** ckDLoqi

**Parents:** aPfw9zP, dYrzLWz, Np4Ajah

**Children:** Z8bgCzz

**Kauro:** Participating in the study is **free** - so there is no cost to you or to your health insurance provider

**Kauro:** If you learn something new about your health as a result of the study, you or your insurance provider will be responsible for the cost of any relevant follow-up testing or medical care from those results

---

**Node ID:** Z8bgCzz

**Parents:** ckDLoqi

**Children:** aBzgDD8

**User Transcript:** Okay

---

**Node ID:** aBzgDD8

**Parents:** Z8bgCzz

**Children:** ApKCtVL

**Kauro:** Just a little more background information for you

**Kauro:** Dr. Eric Vilain is the person responsible for this research study at UCI. This study is paid for by The Human Genome Research Institute (part of the National Institutes of Health or NIH)

---

**Node ID:** ApKCtVL

**Parents:** aBzgDD8

**Children:** 8XuWfkg, UXhKLb9

**Kauro:** We are recruiting patients from all over the country, with all kinds of backgrounds and experiences with genetic testing

---

**Node ID:** 8XuWfkg

**Parents:** ApKCtVL

**Children:** M2fEy3b

**User Transcript:** Makes sense

---

**Node ID:** M2fEy3b

**Parents:** Gs6eCRA, 8XuWfkg

**Children:** Bq68p3L, HQwx7B3

**Kauro:** It’s important to understand that research testing like this does not replace clinical testing.

**Kauro:** Whether you join the PMGRC research study or not, you should always continue getting medical advice from your doctor, which might include other options for genetic testing

**Kauro:** You might want to discuss this study with your family and anyone else you trust before making your decision

---

**Node ID:** Bq68p3L

**Parents:** M2fEy3b

**Children:** 85NXhpr

**User Transcript:** I definitely will

---

**Node ID:** 85NXhpr

**Parents:** YyMv36M, Bq68p3L

**Children:** 2GVi3tK

**Kauro:** So! Let’s talk about what will happen when someone enrolls in this study

---

**Node ID:** 2GVi3tK

**Parents:** 85NXhpr

**Children:** FWCoeMN

**User Transcript:** Yes, please!

---

**Node ID:** FWCoeMN

**Parents:** 2GVi3tK

**Children:** 7SXetE5

**Kauro:** At the end of this chat (or after talking to a study team member), if you decide that you do want to join, you will sign an electronic consent form to officially enroll as a participant.

**Kauro:** Each participant will be assigned a unique study ID number.

**Kauro:** This unique study ID will be used to label your study samples and data (instead of using your name or other identifying information)

---

**Node ID:** 7SXetE5

**Parents:** FWCoeMN

**Children:** LUuSmzi

**User Transcript:** That’s reassuring

---

**Node ID:** LUuSmzi

**Parents:** 7SXetE5

**Children:** dM5aFtN, XxZNtXo, 6wCXYUp

**Kauro:** Once enrolled, you will be asked to provide some medical information and a detailed family health history

**Kauro:** The medical information we collect may include things like doctors’ notes, current or previous diagnoses, and past test results (both genetic and non-genetic)

**Kauro:** If you had previous genetic testing (such as whole genome sequencing), you may be asked to provide the data from this testing

---

**Node ID:** dM5aFtN

**Parents:** LUuSmzi

**Children:** i83McKa

**User Transcript:** Sounds easy enough

---

**Node ID:** i83McKa

**Parents:** dM5aFtN, eX5Kmqt, AJ5y8rf

**Children:** hcVjm7T

**Kauro:** Please be aware that the information we collect will not be provided back to you directly

---

**Node ID:** hcVjm7T

**Parents:** i83McKa

**Children:** Af3kSyQ

**User Transcript:** Okay

---

**Node ID:** Af3kSyQ

**Parents:** hcVjm7T

**Children:** kmCfuJA, 832xEwo, i6bA6Jp

**Kauro:** Once enrolled, you may be asked to provide a tissue sample for genetic testing. This sample could be blood, saliva (spit), or cheek swab (also called “buccal swab”).

**Kauro:** However, not every PMGRC participant will need to provide a sample

---

**Node ID:** kmCfuJA

**Parents:** Af3kSyQ

**Children:** QdRbbvT

**User Transcript:** I understand

---

**Node ID:** QdRbbvT

**Parents:** is9TJxK, 9aApxUq, kmCfuJA

**Children:** jo5VEXs

**Kauro:** And that’s pretty much it! There are no other required interactions

---

**Node ID:** jo5VEXs

**Parents:** QdRbbvT

**Children:** Fok52cR

**User Transcript:** So what happens next?

---

**Node ID:** Fok52cR

**Parents:** jo5VEXs

**Children:** DQsLsQj, dFniP5t

**Kauro:** Once you’ve enrolled and provided the requested information and/or samples, a team of study investigators will review your case history and genetic results

**Kauro:** This will help them decide what research tests will be performed on your samples and data

---

**Node ID:** DQsLsQj

**Parents:** Fok52cR

**Children:** 2DC2FU2

**User Transcript:** That’s easy!

---

**Node ID:** 2DC2FU2

**Parents:** dJioePa, DQsLsQj

**Children:** fjogsGG

**Kauro:** Our PMGRC study staff might contact you to request additional samples or more information from you

---

**Node ID:** fjogsGG

**Parents:** 2DC2FU2

**Children:** 53ZCpC3

**User Transcript:** What if I don’t want to provide additional samples or information?

---

**Node ID:** 53ZCpC3

**Parents:** fjogsGG

**Children:** P32EkeA

**Kauro:** You can decline these additional requests at any time and still be part of the study

---

**Node ID:** P32EkeA

**Parents:** 53ZCpC3

**Children:** 2kEyysx

**User Transcript:** Who has access to the study information?

---

**Node ID:** 2kEyysx

**Parents:** P32EkeA

**Children:** SS9NdGi

**Kauro:** As we mentioned before, the samples and data we gather from you will be labeled only with your unique study ID number. This means that your data and samples are de-identified

**Kauro:** Only a few members of the study team will be able to link your study number to your name

**Kauro:** Other collaborators (such as a commercial lab or a mobile blood draw company) would only have your name and identifiers if you used that company for previous genetic testing or blood collection

---

**Node ID:** SS9NdGi

**Parents:** 2kEyysx

**Children:** dZRNUod

**User Transcript:** That’s good to know

---

**Node ID:** dZRNUod

**Parents:** SS9NdGi

**Children:** 4hnYNnR

**Kauro:** Our PMGRC study is one of 5 Genomics Research Centers. We collaborate with the other 4 centers and might share your genetic information and/or health information with their experts. Any information we share will be de-identified, so these centers will not have access to your name or other personal information that directly identifies you

**Kauro:** This study will also send de-identified data to a National Institutes of Health (NIH)-designated data repository. This repository may be accessed by other researchers around the world

---

**Node ID:** 4hnYNnR

**Parents:** dZRNUod

**Children:** RqztaTu

**User Transcript:** Could my information be used anywhere else?

---

**Node ID:** RqztaTu

**Parents:** 4hnYNnR

**Children:** Nki2gNX

**Kauro:** The results of this research may be presented at scientific meetings or in scientific publications

**Kauro:** You will not be personally identified and no identifiable photos will be published without your specific consent

---

**Node ID:** Nki2gNX

**Parents:** RqztaTu

**Children:** 5vshPHV

**User Transcript:** Okay, that’s good to know

---

**Node ID:** 5vshPHV

**Parents:** Nki2gNX

**Children:** UH32G43

**Kauro:** Your de-identified **genetic information** and **health information** could be used for future research studies or given to another investigator for future research studies without your additional informed consent.

**Kauro:** This de-identified information may be used for many different research studies, for many years in the future.

**Kauro:** If you enroll in the study, you will have the option to let us know if you want to allow your **samples** to be used for other research beyond this study.

**Kauro:** Otherwise, your samples will remain at UCI for use in this study only

---

**Node ID:** UH32G43

**Parents:** 5vshPHV

**Children:** W6x9XW5

**User Transcript:** Good to know!

---

**Node ID:** W6x9XW5

**Parents:** UH32G43

**Children:** ieCqQsR

**Kauro:** So far we've been talking about the ways that de-identified information may be used or shared, but you might be wondering whether identifiable information will ever be shared.

**Kauro:** Your identifiable personal information will not be given to anyone else unless you give your permission in writing first, with a few exceptions that I will explain in a moment.

**Kauro:** The study team is careful with the information we gather for the study, and we will only share it with authorized members of the study team or people who need to review the study information.

---

**Node ID:** ieCqQsR

**Parents:** W6x9XW5

**Children:** 5aJHekM

**User Transcript:** Who might have a need to review the study information?

---

**Node ID:** 5aJHekM

**Parents:** ieCqQsR

**Children:** 4pcbkEp

**Kauro:** There are some third parties such as government agencies or other groups within UCI that may check records that identify you without your permission

**Kauro:** They might look at the study records and your medical records to make sure we are following the law and protecting the people in the study and to make sure our study results are correct

**Kauro:** Some agencies or groups who might see these records are:

- The Department of Health and Human Services Office of Human Research Protections
- The National Human Genome Research Institute (part of the NIH)
- The UCI Medical Center Institutional Review Board (the ethics board that reviewed and approved this research study)
- The Office for the Protection of Human Subjects

---

**Node ID:** 4pcbkEp

**Parents:** 5aJHekM

**Children:** 4SVWLG2

**User Transcript:** Okay, that makes sense

---

**Node ID:** 4SVWLG2

**Parents:** 4SVWLG2

**Children:** eq8AgVx

**Kauro:** You may have heard of HIPAA before, but we want to make sure you know the details and how it applies to your participation in this study

---

**Node ID:** eq8AgVx

**Parents:** 4SVWLG2

**Children:** 3WrbYFW

**User Transcript:** Isn't that something I sign at my doctor's office?

---

**Node ID:** 3WrbYFW

**Parents:** eq8AgVx

**Children:** DB7z6ev

**Kauro:** Yes, you have probably reviewed the HIPAA information at medical visits

**Kauro:** HIPAA is a privacy law that protects your individually identifiable health information (also called "Protected Health Information" or "PHI")

**Kauro:** The **HIPAA** law also describes how information about you may be used or shared if you are in a research study

---

**Node ID:** DB7z6ev

**Parents:** 3WrbYFW

**Children:** QWBbjLo

**User Transcript:** That makes sense

---

**Node ID:** QWBbjLo

**Parents:** DB7z6ev

**Children:** MpZMNKu

**Kauro:** Under HIPAA, you need to sign an agreement in order for researchers to use or share your PHI for research purposes

**Kauro:** I can provide you with a link to UCI's HIPAA Information Sheet at the end of our chat. You can read the complete HIPAA authorization in the consent document

---

**Node ID:** MpZMNKu

**Parents:** QWBbjLo

**Children:** 63uUQep

**User Transcript:** 👍

---

**Node ID:** 63uUQep

**Parents:** MpZMNKu

**Children:** hqiSxfQ

**Kauro:** It is important that you read this carefully and ask a member of the research team to explain anything you do not understand

---

**Node ID:** hqiSxfQ

**Parents:** 63uUQep

**Children:** bTii8uW

**User Transcript:** Can you guarantee that my information will be kept private?

---

**Node ID:** bTii8uW

**Parents:** hqiSxfQ

**Children:** fyZsjfV, NP5jatP

**Kauro:** We will make every effort to keep your information private, but no one's privacy can be totally guaranteed

**Kauro:** To help protect your privacy, the research team has obtained a **Certificate of Confidentiality** from the Department of Health and Human Services (DHHS)

**Kauro:** With this certificate, the investigators cannot be forced (for example, by a court order or subpoena) to give information that may identify you in any federal, state or local civil or criminal court, or in any administrative, legislative, or other proceeding

---

**Node ID:** fyZsjfV

**Parents:** bTii8uW

**Children:** cgwAN3K

**User Transcript:** Good to know

---

**Node ID:** cgwAN3K

**Parents:** QndKtpj, fyZsjfV

**Children:** coJCK6d

**Kauro:** Let's talk about what types of results this research could find

---

**Node ID:** coJCK6d

**Parents:** cgwAN3K

**Children:** UfYVcme

**User Transcript:** Yes!

---

**Node ID:** UfYVcme

**Parents:** coJCK6d

**Children:** AAoupYe

**Kauro:** If your samples and/or data are studied as part of this research, there are different types of results that could be returned to you

**Kauro:** Like we mentioned earlier, you are being referred to this study because you or your family member has a known or suspected genetic condition based on specific health concerns or symptoms

---

**Node ID:** AAoupYe

**Parents:** UfYVcme

**Children:** L82MnJD

**User Transcript:** Right

---

**Node ID:** L82MnJD

**Parents:** AAoupYe

**Children:** 9btMpEf

**Kauro:** This PMGRC study will try to find genetic changes that may be related to those issues

**Kauro:** These results are called **primary findings**, and they often allow the team to make a diagnosis

**Kauro:** Later in this chat, you will get to choose whether you want to be told about these primary findings

---

**Node ID:** 9btMpEf

**Parents:** L82MnJD

**Children:** YJ9BvJu

**User Transcript:** Great!

---

**Node ID:** YJ9BvJu

**Parents:** 9btMpEf

**Children:** XW2AGZ2

**Kauro:** Sometimes the study may find genetic variants that are not related to the patient’s current known health concerns

**Kauro:** These results are called **secondary findings**. Secondary findings are considered **medically actionable**, meaning if you are found to have one of these findings on your testing, then you or your healthcare provider can take action to reduce your risk of developing the health concerns or to better treat or manage the health concerns

---

**Node ID:** XW2AGZ2

**Parents:** YJ9BvJu

**Children:** PLHGemU

**User Transcript:** What kind of findings are medically actionable?

---

**Node ID:** PLHGemU

**Parents:** XW2AGZ2

**Children:** mC7SWtw

**Kauro:** Some examples of medically actionable secondary findings may be genetic variants that cause increased risk for things like heart disease or cancer

**Kauro:** Secondary findings are often related to adult-onset health risks, but it is possible that we could discover a secondary finding that relates to health issues in childhood

---

**Node ID:** mC7SWtw

**Parents:** PLHGemU

**Children:** Et3pvov

**User Transcript:** Okay, I understand

---

**Node ID:** Et3pvov

**Parents:** mC7SWtw

**Children:** Py4Sw9s

**Kauro:** Knowing about a medically actionable genetic variant can help your doctors to adjust your medical care plan

**Kauro:** This can include things like avoiding certain medications or having more frequent checkups with your doctor

---

**Node ID:** Py4Sw9s

**Parents:** Et3pvov

**Children:** Qakavzg

**User Transcript:** That’s good to know

---

**Node ID:** Qakavzg

**Parents:** Py4Sw9s

**Children:** n2xeLXt

**Kauro:** The study could also discover secondary findings that are **not medically actionable**, meaning they could be related to an increased risk for certain diseases with no known treatments or interventions

---

**Node ID:** n2xeLXt

**Parents:** Qakavzg

**Children:** X5VskxR

**User Transcript:** I'll keep that in mind

---

**Node ID:** X5VskxR

**Parents:** n2xeLXt

**Children:** 78Ucru5

**Kauro:** It's important to note that our research team will not be analyzing every sample for secondary findings, but it is possible that we will identify secondary findings in the course of the study

---

**Node ID:** 78Ucru5

**Parents:** X5VskxR

**Children:** NETsCwx

**User Transcript:** What if I don't want to know about health risks from secondary findings?

---

**Node ID:** NETsCwx

**Parents:** 78Ucru5

**Children:** DqZpVap

**Kauro:** That's perfectly fine

**Kauro:** If you do decide to participate in the study, you will get to decide if you want to receive any possible research results. You will decide if you want to receive everything or receive only some types of results and not others

---

**Node ID:** DqZpVap

**Parents:** NETsCwx

**Children:** gAkBYPB

**User Transcript:** That's good to know

---

**Node ID:** gAkBYPB

**Parents:** DqZpVap

**Children:** gabQsoo

**Kauro:** Sometimes genetic testing can find unexpected or surprising results that aren't related to your health

**Kauro:** For example, sometimes testing can reveal that a child's parent is not actually their biological parent or that two parents are related to each other by blood

**Kauro:** These types of unexpected results are called **incidental findings**

---

**Node ID:** gabQsoo

**Parents:** gAkBYPB

**Children:** aYfinQF

**User Transcript:** Oh, okay

---

**Node ID:** aYfinQF

**Parents:** gabQsoo

**Children:** PF2Sqz8

**Kauro:** We do not plan to go looking for these types of incidental findings and we don't plan to tell you about them

**Kauro:** But it is possible that you could figure out these incidental results based on other genetic test results we may share with you

---

**Node ID:** PF2Sqz8

**Parents:** aYfinQF

**Children:** gXJJXUo

**User Transcript:** I understand

---

**Node ID:** gXJJXUo

**Parents:** PF2Sqz8

**Children:** RSiG59t, gEQnvho

**Kauro:** If the study team finds a genetic change that relates to your health concerns, we will tell you about it if you have opted to receive that kind of research result

**Kauro:** Because the results from our study are research results and not clinical results, we will not provide you with an official clinical report

**Kauro:** Any research findings will need to be **confirmed in a certified clinical lab** before they can be added to your medical record

---

**Node ID:** RSiG59t

**Parents:** gXJJXUo, LUkpmdJ

**Children:** 2MNY2vm

**User Transcript:** Who will pay for that additional testing?

---

**Node ID:** 2MNY2vm

**Parents:** RSiG59t

**Children:** kkG2ixC, gEQnvho

**Kauro:** You or your insurance company will have to pay for those additional services

---

**Node ID:** kkG2ixC

**Parents:** 2MNY2vm

**Children:** EJkzrsP

**User Transcript:** Got it

---

**Node ID:** EJkzrsP

**Parents:** YjX3aRS, kkG2ixC

**Children:** Y3WMAjr

**Kauro:** Just so you know, we expect this PMGRC study to last at least until 2026, but you can choose to stop participating at any time

**Kauro:** After signing the Consent/Authorization, you can change your mind and either revoke or withdraw your authorization in the future

---

**Node ID:** Y3WMAjr

**Parents:** EJkzrsP

**Children:** AbjKtPR

**User Transcript:** Good to know!

---

**Node ID:** AbjKtPR

**Parents:** Y3WMAjr

**Children:** MamuHnC

**Kauro:** To revoke the authorization, you must send a written letter to the Principal Investigator to inform him of your decision

**Kauro:** Eric Vilain, MD, PhD
University of California, Irvine (UCI)
Department of Pediatrics
1003 Health Sciences Rd, Suite 308
Irvine, CA 92617

**Kauro:** If you revoke this Authorization, researchers may only use and disclose the PHI that was collected for this research study before you revoked the authorization

**Kauro:** If you change your mind and withdraw the authorization, you will not be allowed to participate in the study any further

---

**Node ID:** MamuHnC

**Parents:** AbjKtPR

**Children:** GKg8Z3M

**User Transcript:** I understand

---

**Node ID:** GKg8Z3M

**Parents:** MamuHnC

**Children:** jdAqu9q

**Kauro:** It is important that you know that anyone who undergoes genetic testing in the United States is protected from certain types of discrimination by a policy called the **Genetic Information Nondiscrimination Act (GINA)**

---

**Node ID:** jdAqu9q

**Parents:** GKg8Z3M

**Children:** VdJaPdJ

**User Transcript:** What areas are protected?

---

**Node ID:** VdJaPdJ

**Parents:** jdAqu9q

**Children:** HE7Z96Y, JkATxFN, Hyni8Jx

**Kauro:** Generally, GINA makes it illegal for health insurance companies, group health insurance plans, and most employers to discriminate against you based on your genetic information

**Kauro:** However, this does not apply to companies with fewer than 15 employees or the United States military

---

**Node ID:** HE7Z96Y

**Parents:** VdJaPdJ, JHvZPbt

**Children:** iyPt6Ye

**User Transcript:** How is health insurance protected?

---

**Node ID:** iyPt6Ye

**Parents:** HE7Z96Y

**Children:** K9gS6Lu, JkATxFN

**Kauro:** Health insurance companies may not request your genetic information that we get from this research

**Kauro:** Health insurance companies may not use your genetic information when deciding whether to insure you or the amount of money they will charge for your plan

---

**Node ID:** K9gS6Lu

**Parents:** iyPt6Ye

**Children:** RpJXYSM

**User Transcript:** I understand

---

**Node ID:** RpJXYSM

**Parents:** HLbqQro, K9gS6Lu, Hyni8Jx

**Children:** EvQQVDh, abqe8JH, htJY7WJ

**Kauro:** It's also very important to know that GINA does not protect you against genetic discrimination by companies that sell life insurance, disability insurance, or long-term care insurance

---

**Node ID:** EvQQVDh

**Parents:** RpJXYSM

**Children:** nk6nfNp

**User Transcript:** What does that mean?

---

**Node ID:** nk6nfNp

**Parents:** EvQQVDh

**Children:** JQkEcaY

**Kauro:** These kinds of additional insurance companies (such as life insurance or disability insurance) might ask you to provide your medical history–including genetic test results–before allowing you to enroll

**Kauro:** Because the genetic testing being done in this study is for research only, the results will not be part of any official medical record until they are confirmed by a clinical lab.

**Kauro:** Some families may choose to enroll in these kinds of insurances for their child or themselves before getting a clinically validated genetic test result

---

**Node ID:** JQkEcaY

**Parents:** nk6nfNp

**Children:** 8NYKg4h

**User Transcript:** Okay

---

**Node ID:** 8NYKg4h

**Parents:** M8MdRDQ, JQkEcaY, htJY7WJ

**Children:** 9s6U2mp

**Kauro:** I’d like to take a moment to give you some info about the **risks and possible discomforts** that may occur as part of participating in this study

**Kauro:** The greatest risks of the study are related to sample collection, the potential for loss of confidentiality of your study information, and possible psychological risks associated with genetic information

**Kauro:** That may sound like a lot, so I will go through each of these with you

---

**Node ID:** 9s6U2mp

**Parents:** 8NYKg4h

**Children:** Co5pFGN

**User Transcript:** Okay, Kauro 👍

---

**Node ID:** Co5pFGN

**Parents:** 9s6U2mp

**Children:** DhAQkSy

**Kauro:** There are some risks related to taking the blood sample, such as discomfort and/or bruising

**Kauro:** Infection, excess bleeding, clotting, or fainting can happen

**Kauro:** There may be minimal discomfort in the cheek if a cheek swab sample is obtained

---

**Node ID:** DhAQkSy

**Parents:** Co5pFGN

**Children:** R87ndyg

**User Transcript:** Okay, I understand

---

**Node ID:** R87ndyg

**Parents:** DhAQkSy

**Children:** YcJnDbx

**Kauro:** The study team will take care to protect the privacy and confidentiality of your study data

**Kauro:** Because your samples and data will be labeled only with a study ID, your personal information (like your name) will only be known to certain authorized study team members

**Kauro:** The team will keep your personal and genetic data in an encrypted secure environment, and be notified immediately if there is a security breach

---

**Node ID:** YcJnDbx

**Parents:** R87ndyg

**Children:** Mz69DPK

**User Transcript:** That’s good to know

---

**Node ID:** Mz69DPK

**Parents:** YcJnDbx

**Children:** UTaGRW8, YDBEbUp

**Kauro:** However, even though your samples and data will not be labeled with your name or other personal information, your genetic information is unique to you

**Kauro:** So it is possible that someone could identify you from your genetic information or learn something about you by looking at your genetic information

**Kauro:** This risk may increase in the future as technologies advance and as more researchers study your genetic information

---

**Node ID:** UTaGRW8

**Parents:** Mz69DPK

**Children:** fsbFZtR

**User Transcript:** Ah, I see

---

**Node ID:** fsbFZtR

**Parents:** YDBEbUp, UTaGRW8

**Children:** D8z5whK

**Kauro:** These risks may also affect members of your family. You may want to discuss your participation in this study with your family and ask the study team about any questions or concerns you may have

---

**Node ID:** D8z5whK

**Parents:** fsbFZtR

**Children:** cMmQUhN

**User Transcript:** Okay, I’ll keep that in mind

---

**Node ID:** cMmQUhN

**Parents:** D8z5whK

**Children:** R6TFjLi

**Kauro:** There can also be some risks associated with learning about genetic results that may be discovered through this study

**Kauro:** Genetic testing can sometimes reveal results that you were not expecting or even things you do not want to know

**Kauro:** It is possible that learning about your genetic results may make you feel distressed or anxious

---

**Node ID:** R6TFjLi

**Parents:** cMmQUhN

**Children:** KbzEchg

**User Transcript:** Ah, okay

---

**Node ID:** KbzEchg

**Parents:** R6TFjLi

**Children:** 3uRFDJg

**Kauro:** These results may be more certain than others

**Kauro:** For example, as we discussed before, we may find a “variant of uncertain significance”, which means that we aren’t sure whether a genetic variant is linked to a specific health concern or not

**Kauro:** Some people may feel anxious or uncomfortable about uncertain results

---

**Node ID:** 3uRFDJg

**Parents:** KbzEchg

**Children:** d4nnpdr

**User Transcript:** Okay

---

**Node ID:** d4nnpdr

**Parents:** 3uRFDJg

**Children:** 9hePugt

**Kauro:** Lastly, results from this study may sometimes give you information about a genetic health issue that does not have a clear medical treatment

**Kauro:** Some people become distressed or anxious to learn about a diagnosis or risk to develop symptoms without a known treatment

**Kauro:** It is up to you to decide if you want to receive any possible genetic results from this study. As we discussed before, you can decide if you want to receive some types of results but not others

---

**Node ID:** 9hePugt

**Parents:** d4nnpdr

**Children:** KX6JCsF

**User Transcript:** That's good to know

---

**Node ID:** KX6JCsF

**Parents:** 9hePugt

**Children:** jey9scg

**Kauro:** Lets also talk about the **possible benefits** of participating in this study

**Kauro:** You should know that there may be no benefit to you for your participation in this study, and you will not be paid for your participation in this study

---

**Node ID:** jey9scg

**Parents:** KX6JCsF

**Children:** h4Luo4M

**User Transcript:** Okay, I understand

---

**Node ID:** h4Luo4M

**Parents:** jey9scg

**Children:** RsJ5kCQ, UynEV5h

**Kauro:** Some study participants may benefit by receiving a new genetic diagnosis or discovering the genetic cause of a known genetic health concern

**Kauro:** This type of information is beneficial because it can help that individual understand more about their health, provide their clinical team some guidance about testing or screening options, and in rare cases, can even help the individual learn about a specific therapy or treatment

---

**Node ID:** RsJ5kCQ

**Parents:** h4Luo4M

**Children:** 7WLks8j

**User Transcript:** Okay, that makes sense

---

**Node ID:** 7WLks8j

**Parents:** UynEV5h, RsJ5kCQ

**Children:** KuX3ekD

**Kauro:** Unfortunately, for many people who have a suspected genetic health concern, a specific diagnosis or cause may never be found

**Kauro:** We do not know if the genetic testing used in this study will be able to help diagnose people who do not yet have a diagnosis

**Kauro:** We hope that through this study we will learn new information and improve our understanding of genetic conditions to help us find diagnoses for people in the future

---

**Node ID:** KuX3ekD

**Parents:** 7WLks8j

**Children:** nYueqrk

**User Transcript:** Okay

---

**Node ID:** nYueqrk

**Parents:** KuX3ekD

**Children:** 5UhK2Hy

**Kauro:** I'm going to give you some contact information, and you can write it down if you would like. ✏️

**Kauro:** If you have any questions about the study or if you get hurt or sick because of something to do with the study, please contact the principal investigator, Dr. Eric Vilain, at 949-824-4090.

---

**Node ID:** 5UhK2Hy

**Parents:** nYueqrk

**Children:** BXUr8TN

**User Transcript:** Thanks for this information

---

**Node ID:** BXUr8TN

**Parents:** 5UhK2Hy

**Children:** QvCduQU

**Kauro:** If you have any questions or concerns about your privacy rights, you may contact the UCI Health Privacy Office at 714-456-3916

**Kauro:** If you have questions or concerns about your rights as a study participant, you can contact UCI's Human Research Protections unit in the Office of Research at 949-824-6068, or by email, or by writing to us at 160 Aldrich Hall, Irvine, CA 92697

---

**Node ID:** QvCduQU

**Parents:** BXUr8TN

**Children:** Dq4hG9k

**User Transcript:** Okay

---

**Node ID:** Dq4hG9k

**Parents:** QvCduQU

**Children:** BKUeKkp, FVdUisi, NsfXTD7, BMeYncH

**Kauro:** Whew! 😅 We just went through a lot of information together

**Kauro:** I've gone through all the topics I need to cover

**Kauro:** Now, it's up to you to decide if you would like to give your consent for yourself or your child to participate in the PMGRC study

---

**Node ID:** BKUeKkp

**Parents:** Dq4hG9k, h9rpY7K, ixSPowJ

**Children:** SZabuK4

**User Transcript:** I am ready to consent!

---

**Node ID:** SZabuK4

**Parents:** BKUeKkp

**Children:** bmorWZo

**Kauro:** Okay, great 😀

**Kauro:** In a moment, I will ask you to make some choices about how you want to participate

**Kauro:** But first, I need to ask you some questions to make sure you understand the basics of participating in the PMGRC study

**Kauro:** And don't worry - if you answer any of these questions incorrectly, I'll help remind you of the right answer

---

**Node ID:** bmorWZo

**Parents:** SZabuK4

**Children:** VWzWNAK

**User Transcript:** Okay - I'm ready

---

**Node ID:** VUipQHn

**Parents:** nFpTPVg

**Children:** JTCaWss

**Kauro:** You got 10 out of 10 questions right! 🥇

**Kauro:** Now we're ready to collect your consent for participation in the study

**Kauro:** We'll start by capturing your enrollment preferences and then we'll ask for your electronic signature

---

**Node ID:** QKNbLGv

**Parents:** JTCaWss

**Children:** gdiDJUS

**Kauro:** We would like to store your research samples in a biobank for future research, which we will describe in a moment

**Kauro:** The samples we store could include genetic material (DNA or RNA), skin or other tissue samples, or cells

**Kauro:** The biobank is maintained by the study team at UCI

**Kauro:** Please let us know how you would like your samples to be used

---

**Node ID:** gdiDJUS

**Parents:** QKNbLGv

**Children:** Psu2bBx

**Form ID:** sample-storage-use-form

**From Text:**

Can we store your samples for purposes of this research study? (Required for participation in the study)

Can we store your samples in the UCI biobank for purposes of unrelated research?

**From Options:**


```
[
  {
    "name": "storeSamplesThisStudy",
    "type": "radio",
    "label": "Study Sample Storage",
    "options": [
      {
        "label": "Yes",
        "value": "yes"
      }
    ]
  },
  {
    "name": "storeSamplesOtherStudies",
    "type": "radio",
    "label": "Other Studies Sample Storage",
    "options": [
      {
        "label": "Yes",
        "value": "yes"
      },
      {
        "label": "No",
        "value": "no"
      }
    ]
  }
]
```


---

**Node ID:** Psu2bBx

**Parents:** gdiDJUS

**Children:** bdmfVYQ

**Kauro:** You should know that you may change your mind at a later time and request that your sample be destroyed

**Kauro:** If you change your mind and want to request that your sample be destroyed, you must send a written request to:

Eric Vilain, MD, PhD
University of California, Irvine (UCI)
Department of Pediatrics
1003 Health Sciences Rd, Suite 308
Irvine, CA 92617

---

**Node ID:** bdmfVYQ

**Parents:** Psu2bBx

**Children:** cmJfBjf

**User Transcript:** That’s good to know

---

**Node ID:** cmJfBjf

**Parents:** bdmfVYQ

**Children:** VZsTdFg

**Kauro:** We would like to store personal health information (PHI) collected from you in this study (such as your name, date of birth, contact information, etc.) in a database for future research

**Kauro:** The database is maintained by the study team at UCI

**Kauro:** Please let us know how you would like your PHI to be stored and used

---

**Node ID:** VZsTdFg

**Parents:** cmJfBjf

**Children:** 4bYChBx

**Form ID:** phi-use-form

**From Text:**

Can we store your PHI in our database for research related to this study? (Required for participation in the study)

Can we use your stored PHI to contact you about future research opportunities that are not related to this study?

**From Options:**

```
[
  {
    "name": "storePhiThisStudy",
    "type": "radio",
    "label": "PHI Storage for Study",
    "options": [
      {
        "label": "Yes",
        "value": "yes"
      }
    ]
  },
  {
    "name": "storePhiOtherStudies",
    "type": "radio",
    "label": "PHI Use for Other Studies",
    "options": [
      {
        "label": "Yes",
        "value": "yes"
      },
      {
        "label": "No",
        "value": "no"
      }
    ]
  }
]
```


---

**Node ID:** 4bYChBx

**Parents:** VZsTdFg

**Children:** SLcyBjQ, GzG3exY

**Kauro:** You may recall that we discussed the types of genetic research results that may be found in this study

**Kauro:** You get to decide which types of results you want to learn about

**Kauro:** Do you remember the different types of results we discussed, like “primary results” and “medically actionable secondary results”?

---

**Node ID:** SLcyBjQ

**Parents:** 4bYChBx

**Children:** QpzocPu

**User Transcript:** Yes, I remember

---

**Node ID:** QpzocPu

**Parents:** hnqNqbi, SLcyBjQ

**Children:** nriTamw

**Kauro:** Please complete the form below to let us know what types of results you would like to receive

---

**Node ID:** nriTamw

**Parents:** QpzocPu

**Children:** knbtYck

**Form ID:** result-return-form

**From Text:**

**From Options:**


```
[
  {
    "name": "rorPrimary",
    "type": "radio",
    "label": "Do you want to learn about primary results that are found?",
    "options": [
      {
        "label": "Yes",
        "value": "yes"
      },
      {
        "label": "No",
        "value": "no"
      }
    ]
  },
  {
    "name": "rorSecondary",
    "type": "radio",
    "label": "Do you also want to learn about medically actionable secondary results?",
    "options": [
      {
        "label": "Yes",
        "value": "yes"
      },
      {
        "label": "No",
        "value": "no"
      }
    ]
  },
  {
```

```
    "name": "rorSecondaryNot",
    "type": "radio",
    "label": "Do you want to learn about secondary results that are NOT
medically actionable?",
    "options": [
      {
        "label": "Yes",
        "value": "yes"
      },
      {
        "label": "No",
        "value": "no"
      }
    ]
  }
]
```


---

**Node ID:** knbtYck

**Parents:** nriTamw

**Children:** AimWGCA

**Kauro:** Great! You have now entered all of your choices for your enrollment

**Kauro:** You are ready to provide your signature to consent!

---

**Node ID:** 5n6QHJ7

**Parents:** AimWGCA

**Children:** Mzh9KYH

**Kauro:** Thank you!

---

**Node ID:** Mzh9KYH

**Parents:** nAaeApX, 5n6QHJ7, VbbeudE

**Children:** KxvpjwE

**Kauro:** We are almost done with the consent process. Just a few more things to tell you.

**Kauro:** Below are links to access some of the documents we discussed during our chat:

**HIPAA information** - https://www.cdc.gov/phlp/publications/topic/hipaa.html

**GINA resources** - https://www.genome.gov/about-genomics/policy-issues/Genetic-Discrimination#gina

**CalGINA information** - http://www.leginfo.ca.gov/pub/11-12/bill/sen/sb_0551-0600/sb_559_bill_20110906_chaptered.pdf

---

**Node ID:** KxvpjwE

**Parents:** Mzh9KYH, Mzke78A

**Children:** W7DeBPU

**User Transcript:** Thank you

---

**Node ID:** W7DeBPU

**Parents:** KxvpjwE

**Children:** TSRVTj5

**Kauro:** After our study staff has finalized the consent paperwork, we will send you a copy of the signed consent documents via a secure email using the email address you provided above

**Kauro:** You will receive a separate email with a transcript of our chat

---

**Node ID:** TSRVTj5

**Parents:** W7DeBPU

**Children:** 39kDMR8

**User Transcript:** Okay, great!

---

**Node ID:** 39kDMR8

**Parents:** TSRVTj5

**Children:** 6iruWjc

**Kauro:** We will also send you an email with a link to an online form where we will collect health information to complete your study enrollment

**Kauro:** Once you are enrolled, we will be in touch to let you know if we need to collect any samples from you or your child

---

**Node ID:** 6iruWjc

**Parents:** 39kDMR8

**Children:** nJgsH7y

**User Transcript:** That sounds good

---

**Node ID:** nJgsH7y

**Parents:** nqDwZrd, 6iruWjc, MNpRjAA, PZYk7Ga

**Children:** BUgU8Tq, nTBdMds

**Kauro:** Remember, if you have any questions or concerns, you can always reach out to our team at

**Kauro:** If you would like to help me improve, please rate your experience and provide feedback!

---

**Node ID:** BUgU8Tq

**Parents:** nJgsH7y

**Children:** YQH834A, aXDDbh2

**User Transcript:** I'd like to leave feedback

---

**Node ID:** YQH834A

**Parents:** BUgU8Tq

**Children:** aXDDbh2

**Kauro:** Great, thanks so much!

---

**Node ID:** aXDDbh2

**Parents:** BUgU8Tq, YQH834A

**Children:** YQHvnrA

**Form ID:** user-feedback-form

**From Text:**

**From Options:**


```
[
  {
    "name": "satisfaction",
    "type": "select",
    "label": "Overall satisfaction with the consent process",
    "options": [
      {
        "label": "Very Satisfied",
        "value": "Very Satisfied"
      },
      {
        "label": "Satisfied",
        "value": "Satisfied"
      },
      {
        "label": "Neutral",
        "value": "Neutral"
      },
      {
        "label": "Dissatisfied",
        "value": "Dissatisfied"
      },
```

```
      {
        "label": "Very Dissatisfied",
        "value": "Very Dissatisfied"
      }
    ],
    "required": true
  },
  {
    "name": "suggestions",
    "type": "textarea",
    "label": "Suggestions for Improvement"
  }
]
```


---

**Node ID:** nTBdMds

**Parents:** nJgsH7y

**Children:** 7PxxtTh

**User Transcript:** No thanks

---

**Node ID:** GzG3exY

**Parents:** 4bYChBx

**Children:** 3iBfwGK

**User Transcript:** No, can you remind me?

---

**Node ID:** 3iBfwGK

**Parents:** GzG3exY

**Children:** WHwSvgC

**Kauro: Primary findings** are genetic changes that may be related to the specific health concern that has been seen in you or your family members

**Kauro:** Primary findings often allow the clinical team to make a diagnosis

---

**Node ID:** WHwSvgC

**Parents:** 3iBfwGK

**Children:** N9JZqfY

**User Transcript:** Okay

---

**Node ID:** N9JZqfY

**Parents:** WHwSvgC

**Children:** Hc525XA

**Kauro:** Sometimes the study may find genetic variants that are not related to a patient’s current known health concerns

**Kauro:** These are called **secondary findings**

**Kauro:** Some of these secondary findings are considered medically actionable, meaning if these genetic changes are known, then you or your healthcare provider can then take action to try to reduce your risk of developing the symptoms or to better treat or manage your healthcare related to the secondary findings

---

**Node ID:** Hc525XA

**Parents:** N9JZqfY

**Children:** GMm2FJg

**User Transcript:** What kind of findings are medically actionable?

---

**Node ID:** GMm2FJg

**Parents:** Hc525XA

**Children:** hnqNqbi

**Kauro:** Some examples of medically actionable secondary findings may be genetic variants that cause increased risk for future health concerns like heart disease or cancer

**Kauro:** Secondary findings are often related to adult-onset health risks. However, it is possible that we could discover a secondary finding that relates to symptoms in childhood

---

**Node ID:** hnqNqbi

**Parents:** GMm2FJg

**Children:** QpzocPu

**User Transcript:** Okay

---

**Node ID:** nbkJ3g7

**Parents:** JTCaWss

**Children:** YkoAEVe

**Kauro:** Now, let’s talk about the next steps for enrolling your child or children

**Kauro:** How many children are you enrolling?

---

**Node ID:** YkoAEVe

**Parents:** nbkJ3g7

**Children:** eDqsGdn, VbbeudE

**Form ID:** num-children-enroll-form

**From Text:**

**From Options:**


```
[
  {
    "name": "numChildrenEnroll",
    "type": "radio",
    "label": "How many children are you enrolling?",
    "options": [
      {
        "label": "1",
        "value": "1"
      },
      {
        "label": "2",
        "value": "2"
      },
      {
        "label": "3",
        "value": "3"
      },
      {
        "label": "4 or more",
        "value": "4"
      }
    ]
  }
]
```


---

**Node ID:** Mzke78A

**Parents:** nAaeApX

**Children:** KxvpjwE

**Kauro:** We are almost done with the consent process. Just a few more things to tell you.

**Kauro:** If you enrolled a child who is 7 years or older, a member of the study team will need to talk to your child briefly to tell them about the study and confirm their interest in participating

**Kauro:** Below are links to access some of the documents we discussed during our chat:

**HIPAA information** - https://www.cdc.gov/phlp/publications/topic/hipaa.html

**GINA resources** - https://www.genome.gov/about-genomics/policy-issues/Genetic-Discrimination#gina

**CalGINA information** - http://www.leginfo.ca.gov/pub/11-12/bill/sen/sb_0551-0600/sb_559_bill_20110906_chaptered.pdf

---

**Node ID:** VbbeudE

**Parents:** YkoAEVe

**Children:** Mzh9KYH

**Kauro:** Okay, great!

**Kauro:** To enroll four or more children, it would be best for you to talk to a member of our study team

**Kauro:** Someone from our team will contact you directly by phone or email within the next 2-3 weeks

**Kauro:** If you have questions now, you can contact us at

---

**Node ID:** G3EXSPz

**Parents:** nFpTPVg

**Children:** DdxFUae, kZ6qj4C

**Kauro:** It seems like you had a little trouble with these questions

**Kauro:** I hope my responses helped clarify.

**Kauro:** Would you like to go through them one more time, or would you like to talk to a member of our study team?

---

**Node ID:** nqDwZrd

**Parents:** DdxFUae

**Children:** nJgsH7y

**Kauro:** Sure thing

**Kauro:** Someone from our study team will contact you directly by phone or email within the next 2-3 weeks

---

**Node ID:** iH6N9fF

**Parents:** nFpTPVg

**Children:** X4CvZHZ

**Kauro:** I am sorry. You did not get all the answers correct.

**Kauro:** To complete your consent, we’d like to connect you to a member of our study team

---

**Node ID:** PZYk7Ga

**Parents:** X4CvZHZ

**Children:** nJgsH7y

**Kauro:** Someone from our team will contact you directly by phone or email within the next 2-3 weeks

---

**Node ID:** Swxbt6c

**Parents:** FVdUisi

**Children:** MNpRjAA

**Kauro:** Okay - that's just fine

---

**Node ID:** MNpRjAA

**Parents:** Swxbt6c

**Children:** nJgsH7y

**User Transcript:** Okay, sounds good!

---

**Node ID:** P8C5vVo

**Parents:** NsfXTD7

**Children:** kiRj7cx

**Kauro:** Okay! Please take all the time you need

**Kauro:** As you are considering things, you can scroll through our chat to review our discussion any time. To come back to the chat, click the link you used to access the chat

**Kauro:** If the link is expired or does not work, the study team can send a new link

---

**Node ID:** kiRj7cx

**Parents:** P8C5vVo

**Children:** ixSPowJ

**User Transcript:** Sounds good

---

**Node ID:** ixSPowJ

**Parents:** kiRj7cx

**Children:** BKUeKkp, FVdUisi

**Kauro:** When you are ready to decide, you can come back and continue this chat

---

**Node ID:** h9rpY7K

**Parents:** BMeYncH

**Children:** BKUeKkp, FVdUisi

**Kauro:** Okay! I can arrange for someone from our study team to contact you directly by phone and/or email within the next 2-3 weeks

**Kauro:** You can also come back here to continue the chat if you have made a decision

---

**Node ID:** UynEV5h

**Parents:** h4Luo4M

**Children:** 7WLks8j

**User Transcript:** That would be great

---

**Node ID:** YDBEbUp

**Parents:** Mz69DPK

**Children:** fsbFZtR

**User Transcript:** That is concerning

---

**Node ID:** abqe8JH

**Parents:** RpJXYSM

**Children:** nB4pVm8

**User Transcript:** Do I need to worry about this?

---

**Node ID:** nB4pVm8

**Parents:** abqe8JH

**Children:** M8MdRDQ

**Kauro:** Because the genetic testing being done in this study is for research only, the results will not be part of any official medical record until they are confirmed by a clinical lab

**Kauro:** Some families may choose to enroll in these kinds of insurances for their child or themselves before getting a clinically-validated genetic test result

---

**Node ID:** M8MdRDQ

**Parents:** nB4pVm8

**Children:** 8NYKg4h

**User Transcript:** Okay

---

**Node ID:** htJY7WJ

**Parents:** RpJXYSM

**Children:** 8NYKg4h

**User Transcript:** Okay

---

**Node ID:** JkATxFN

**Parents:** VdJaPdJ, iyPt6Ye

**Children:** JHvZPbt

**User Transcript:** How are employers involved?

---

**Node ID:** JHvZPbt

**Parents:** JkATxFN

**Children:** HLbqQro, HE7Z96Y

**Kauro:** Employers may not use your genetic information that we get from this research when deciding to hire, promote, or fire you

---

**Node ID:** HLbqQro

**Parents:** JHvZPbt

**Children:** RpJXYSM

**User Transcript:** I understand

---

**Node ID:** Hyni8Jx

**Parents:** VdJaPdJ

**Children:** RpJXYSM

**User Transcript:** I understand

---

**Node ID:** gEQnvho

**Parents:** gXJJXUo, 2MNY2vm

**Children:** LUkpmdJ

**User Transcript:** What does "confirmed in a clinical lab" mean?

---

**Node ID:** LUkpmdJ

**Parents:** gEQnvho

**Children:** YjX3aRS, RSiG59t

**Kauro:** Most tests done on samples in research studies are **only for research** and have no clear meaning for healthcare

**Kauro:** If we think that there are research results that might have meaning for you or your family, these tests should be re-done by a certified clinical laboratory to confirm them

**Kauro:** Results will only be placed in your medical record once they have been clinically confirmed

---

**Node ID:** YjX3aRS

**Parents:** LUkpmdJ

**Children:** EJkzrsP

**User Transcript:** I understand

---

**Node ID:** NP5jatP

**Parents:** bTii8uW

**Children:** 9SsWRKf

**User Transcript:** What if I tell people I'm part of this study?

---

**Node ID:** 9SsWRKf

**Parents:** NP5jatP

**Children:** QndKtpj

**Kauro:** It is important that you know that a Certificate of Confidentiality does not stop you or a member of your family from voluntarily giving information to others about you or about taking part in this research

**Kauro:** You should also know that if an insurer or employer learns about your participation and you give them permission to receive research information about you, we cannot use the Certificate of Confidentiality to keep your information private from them

**Kauro:** This means that you must also actively protect your own privacy

---

**Node ID:** QndKtpj

**Parents:** 9SsWRKf

**Children:** cgwAN3K

**User Transcript:** Got it

---

**Node ID:** dFniP5t

**Parents:** Fok52cR

**Children:** dJioePa

**User Transcript:** What kinds of research tests?

---

**Node ID:** dJioePa

**Parents:** dFniP5t

**Children:** 2DC2FU2

**Kauro:** Your sample may be used for a variety of genetic testing including, but not limited to: Whole Genome Sequencing, Optical Mapping, and RNA sequencing.

**Kauro:** These are different ways of looking at your genetic material

---

**Node ID:** 832xEwo

**Parents:** Af3kSyQ, CFMdbEP

**Children:** jzxix3y

**User Transcript:** Where will my blood be drawn?

---

**Node ID:** jzxix3y

**Parents:** 832xEwo

**Children:** 9aApxUq, i6bA6Jp

**Kauro:** There are a few options

**Kauro:** If you are having clinical blood drawn for another reason, an extra sample may be collected for this study at that time

**Kauro:** You could also schedule a blood draw at the UCI research unit or ask if our study team is able to arrange for a mobile phlebotomy company to come to your home for a blood draw

---

**Node ID:** 9aApxUq

**Parents:** jzxix3y

**Children:** QdRbbvT

**User Transcript:** So there are options. Got it

---

**Node ID:** i6bA6Jp

**Parents:** Af3kSyQ, jzxix3y

**Children:** CFMdbEP

**User Transcript:** What’s a buccal swab?

---

**Node ID:** CFMdbEP

**Parents:** i6bA6Jp

**Children:** is9TJxK, 832xEwo

**Kauro:** A “buccal swab” is like a large Q-tip used for swabbing the inside of the cheeks.

**Kauro:** If you provide a buccal sample, these will be collected at home with a kit provided to you with instructions for collection at home and mailing back. Sometimes your doctor at UCI can help you collect your buccal swab

---

**Node ID:** is9TJxK

**Parents:** CFMdbEP

**Children:** QdRbbvT

**User Transcript:** Cool!

---

**Node ID:** XxZNtXo

**Parents:** LUuSmzi

**Children:** nUA9owA

**User Transcript:** Where do I get my doctor's notes?

---

**Node ID:** nUA9owA

**Parents:** XxZNtXo, SvnYgFC

**Children:** AJ5y8rf, 6wCXYUp

**Kauro:** Most doctor's offices and hospitals have a "Medical Records" office or a patient portal where notes and test results can be sent directly to the patient.

**Kauro:** Our study staff can help you take the next steps, if necessary

---

**Node ID:** AJ5y8rf

**Parents:** nUA9owA

**Children:** i83McKa

**User Transcript:** I see

---

**Node ID:** 6wCXYUp

**Parents:** LUuSmzi, nUA9owA

**Children:** ikGZqyF

**User Transcript:** Where do I get my genetic testing data?

---

**Node ID:** ikGZqyF

**Parents:** 6wCXYUp

**Children:** eX5Kmqt, SvnYgFC

**Kauro:** Usually, you will provide a signed medical release to us, which we will send to the lab that did the original testing or to your doctor's office. Our study team can help you take the next steps, if necessary

---

**Node ID:** eX5Kmqt

**Parents:** ikGZqyF

**Children:** i83McKa

**User Transcript:** Okay

---

**Node ID:** SvnYgFC

**Parents:** ikGZqyF

**Children:** nUA9owA

**User Transcript:** Where do I get my doctor's notes?

---

**Node ID:** HQwx7B3

**Parents:** M2fEy3b

**Children:** 2SpZFSY

**User Transcript:** Who should I talk to?

---

**Node ID:** 2SpZFSY

**Parents:** HQwx7B3

**Children:** YyMv36M

**Kauro:** Some people you could talk to about your participation in this study include your family members, your doctor, or someone you trust to help you make decisions

---

**Node ID:** YyMv36M

**Parents:** 2SpZFSY

**Children:** 85NXhpr

**User Transcript:** Okay

---

**Node ID:** UXhKLb9

**Parents:** ApKCtVL

**Children:** Gb4EZXT

**User Transcript:** How many people will be involved?

---

**Node ID:** Gb4EZXT

**Parents:** UXhKLb9

**Children:** Gs6eCRA

**Kauro:** We are hoping to enroll up to 4,200 people.

**Kauro:** We expect half to be patients and family members recruited at UCI and half to be referred from outside of UCI

---

**Node ID:** Gs6eCRA

**Parents:** Gb4EZXT

**Children:** M2fEy3b

**User Transcript:** Got it

---

**Node ID:** HUAk3zG

**Parents:** NHPvQRe

**Children:** Gufd5fp

**User Transcript:** What if I am a student or employee at UCI?

---

**Node ID:** Gufd5fp

**Parents:** 8BCaaXA, HUAk3zG

**Children:** aPfw9zP, k9yQNWE

**Kauro:** If you are an employee or student (undergraduate, graduate, medical) in training at UCI, your decision to participate or not participate will not affect your employment or academic standing

---

**Node ID:** aPfw9zP

**Parents:** Gufd5fp

**Children:** ckDLoqi

**User Transcript:** Great!

---

**Node ID:** k9yQNWE

**Parents:** Gufd5fp

**Children:** aJ5R3Ko

**User Transcript:** How do I let you know if I no longer want to be in the study?

---

**Node ID:** aJ5R3Ko

**Parents:** Ua23Tdf, k9yQNWE

**Children:** dYrzLWz, 8BCaaXA

**Kauro:** To withdraw from the study, you must send a written letter to the Principal Investigator (Dr. Eric Vilain) to inform him of your decision

---

**Node ID:** dYrzLWz

**Parents:** aJ5R3Ko

**Children:** ckDLoqi

**User Transcript:** Sounds good!

---

**Node ID:** 8BCaaXA

**Parents:** aJ5R3Ko

**Children:** Gufd5fp

**User Transcript:** What if I am a student or employee at UCI?

---

**Node ID:** Ua23Tdf

**Parents:** NHPvQRe

**Children:** aJ5R3Ko

**User Transcript:** How do I let you know if I no longer want to be in the study?

---

**Node ID:** LbgZ4YK

**Parents:** SgDDtr8

**Children:** ZjLbVa4

**User Transcript:** Tell me more about the different types of genetic test results?

---

**Node ID:** ZjLbVa4

**Parents:** LbgZ4YK

**Children:** h2xWJaa

**Kauro:** Of course!

**Kauro:** When genetic testing is done, we are usually looking for a genetic variant or variants to explain the patient’s health concerns

**Kauro:** When the testing finds an answer, meaning a genetic variant(s) that explains your health concerns is identified, we call this a “positive” result

---

**Node ID:** h2xWJaa

**Parents:** ZjLbVa4

**Children:** WAkoAEV

**User Transcript:** Okay

---

**Node ID:** WAkoAEV

**Parents:** h2xWJaa

**Children:** DKTqMXc

[redacted]

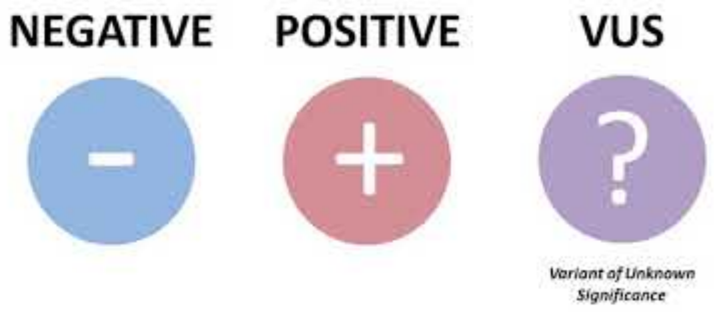


[redacted]

**Kauro:** Video link: https://www.youtube-nocookie.com/embed/HbQTI2pDmSs

**Kauro:** If the testing does NOT find any variants that could explain your health concern, we call this a “negative” result

**Kauro:** Sometimes, the testing can find something that we do not fully understand

**Kauro:** When a variant is found, but we aren’t sure whether it is linked to a symptom, we call this a “variant of uncertain significance” or VUS

**Kauro:** Here is a brief video to review the types of genetic results we just discussed:

---

**Node ID:** DKTqMXc

**Parents:** WAkoAEV

**Children:** XKshQvX

**User Transcript:** Ah, okay

---

**Node ID:** 9cuqxwT

**Parents:** jczufDn

**Children:** nVUpS6V

**User Transcript:** Had genetic testing

---

**Node ID:** nVUpS6V

**Parents:** 9cuqxwT

**Children:** kYjpLjd

**Kauro:** So you’ve had some experience. That’s good!

---

**Node ID:** kYjpLjd

**Parents:** nVUpS6V

**Children:** FiRFHGF

**Kauro:** One goal for our PMGRC study is to discovery new causes of health problems, like finding a new gene or new ways that differences in our genes can cause health concerns

**Kauro:** If genetic testing didn't find an answer before, our research might be able to find something new

---

**Node ID:** iVCEBuL

**Parents:** EE6kxBa

**Children:** 8Z6qtgu

**User Transcript:** That makes sense

---

**Node ID:** 9ZmfvBQ

**Parents:** 45kz8CE

**Children:** BF3k6W7

**User Transcript:** Tell me more

---

**Node ID:** QXNa2cz

**Parents:** Ey8gJrc, WnXoffX

**Children:** abx8PPP

**User Transcript:** Let's keep it simple

---

**Node ID:** abx8PPP

**Parents:** QXNa2cz

**Children:** jUSVMUz, W5Gtph5

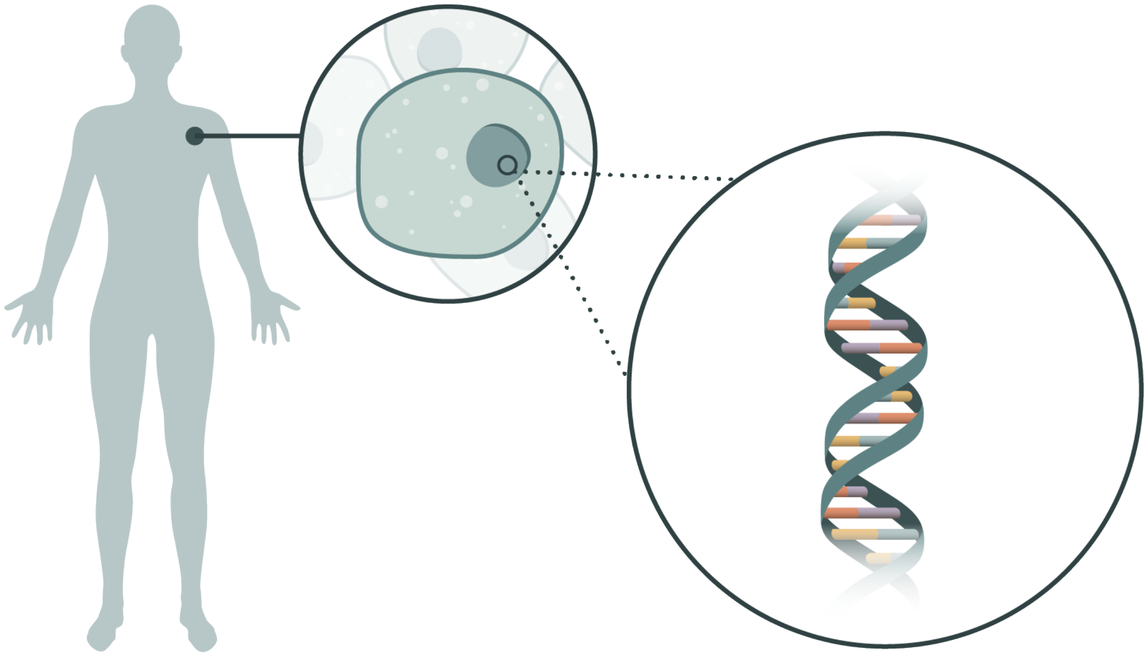

**Kauro:** Okay, sounds good

**Kauro:** You may already know that your body is made of trillions of cells. DNA provides the instructions to tell our bodies how to grow and develop

---

**Node ID:** jUSVMUz

**Parents:** abx8PPP

**Children:** 2wRsFE8

**User Transcript:** Okay

---

**Node ID:** 2wRsFE8

**Parents:** W5Gtph5, jUSVMUz

**Children:** VNSy2sy

**Kauro:** Genes are a big part of what makes you who you are

**Kauro:** In your cells, each gene provides the instructions for molecules called RNA, and each RNA makes a different protein

---

**Node ID:** VNSy2sy

**Parents:** 2wRsFE8

**Children:** 6cFB5vN

**User Transcript:** That’s interesting

---

**Node ID:** 6cFB5vN

**Parents:** VNSy2sy

**Children:** eo5YNAs, fFHd2uw

**Kauro:** Your body uses proteins to do many important things

**Kauro:** For example, some proteins build our tissues, like muscles and nerves. Other proteins help our body break down food and make energy

---

**Node ID:** eo5YNAs

**Parents:** 6cFB5vN

**Children:** jbTfjLx

**User Transcript:** Okay

---

**Node ID:** jbTfjLx

**Parents:** fFHd2uw, eo5YNAs

**Children:** TXdMNWZ

**Kauro:** We all have small differences in our genes, which we call "variants"

---

**Node ID:** TXdMNWZ

**Parents:** jbTfjLx

**Children:** PiTC5fc

**User Transcript:** I see

---

**Node ID:** PiTC5fc

**Parents:** TXdMNWZ

**Children:** Ti3ZavP

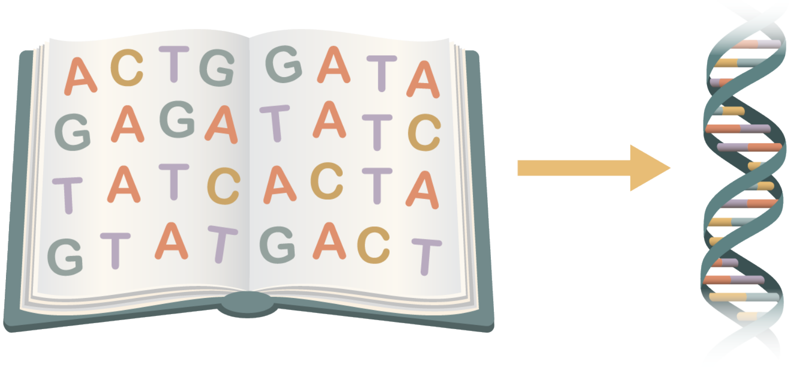


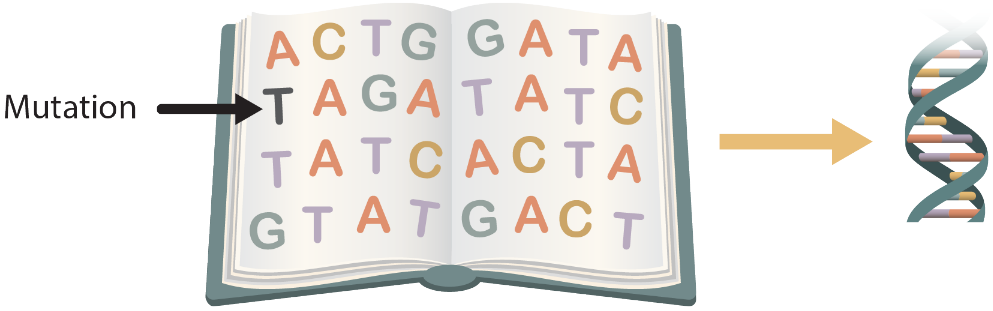


**Kauro:** When a gene has a variant it gives the body different instructions. It’s like changing an ingredient in a recipe

---

**Node ID:** Ti3ZavP

**Parents:** PiTC5fc

**Children:** EE6kxBa

**User Transcript:** Got it

---

**Node ID:** fFHd2uw

**Parents:** 6cFB5vN

**Children:** jbTfjLx

**User Transcript:** That’s cool!

---

**Node ID:** W5Gtph5

**Parents:** abx8PPP

**Children:** 2wRsFE8

**User Transcript:** This sounds familiar

---

**Node ID:** WnXoffX

**Parents:** AXCfXjh, 4iXP7YP, Sj26JXx

**Children:** KR89wri, QXNa2cz

**Kauro:** Now, let’s take a moment to talk a little bit about genetics.

**Kauro:** Do you want to keep it simple or get a little more science-y?

---

**Node ID:** VDz6Yx8

**Parents:** RMb2hrx

**Children:** 3jPsush

**User Transcript:** One child

---

**Node ID:** 3jPsush

**Parents:** VDz6Yx8, 5ZrtE33

**Children:** GkgFmXm

**Kauro:** Okay, can you please select the ages of your children?

---

**Node ID:** Sj26JXx

**Parents:** DuLoHj9, fmNaDgk, WtmwJYx, mGbcBU9, 5hX84fN

**Children:** WnXoffX

**Kauro:** You’re welcome 😀

---

**Node ID:** 5ZrtE33

**Parents:** RMb2hrx

**Children:** 3jPsush

**User Transcript:** More than one child

---

**Node ID:** CAwKwCW

**Parents:** CPf9CCz

**Children:** USfk36C

**User Transcript:** I understand

---

**Node ID:** Bkyp7hD

**Parents:** FADCStr

**Children:** QT8Mx3R

**User Transcript:** Why only those related by blood?

---

**Node ID:** QT8Mx3R

**Parents:** Bkyp7hD

**Children:** RXBDyqd

**Kauro:** Having genetic data from blood relatives can help us understand genetic findings

**Kauro:** For example, if we find a genetic change in a person who has specific medical concern, like heart disease, it can be helpful to know if this genetic change was passed down through the family to identify other individuals at risk

---

**Node ID:** RXBDyqd

**Parents:** QT8Mx3R

**Children:** KMirb2i

**User Transcript:** Ah, that makes sense

---

**Node ID:** BxD4wrq

**Parents:** S2nH9zZ

**Children:** jbs93Hc

**User Transcript:** I want to pick up this chat with Kauro sometime later

---

**Node ID:** jbs93Hc

**Parents:** BxD4wrq

**Children:** 87q8gWG

**Kauro:** Okay! No problem

**Kauro:** You can use the link in your text message to come back at any time

**Kauro:** Let me know when you are ready to chat

---

**Node ID:** 87q8gWG

**Parents:** jbs93Hc

**Children:** Q8yaxQc

**User Transcript:** I’m ready to chat now

---

**Node ID:** iLpXdAS

**Parents:** oS9m8Ua

**Children:** a6JKRdh

**User Transcript:** I understand

---

#### Enroll Form

**Node ID:** b5nYNf6

**Parents:** USfk36C

**Children:** eca6cQF, axvGUBt, RMb2hrx

**Form ID:** checkbox-form

**From Text:**

**From Options:**


```
[
  {
    "name": "myself",
    "label": "Myself",
    "value": "Myself",
    "id_value": "eca6cQF"
  },
  {
    "name": "myChildChildren",
    "label": "My child/children",
    "value": "My child/children",
    "id_value": "RMb2hrx"
  },
  {
    "name": "childOtherParent",
    "label": "My child\u2019s other parent",
    "value": "My child\u2019s other parent",
    "id_value": "axvGUBt"
  },
  {
    "name": "adultFamilyMember",
    "label": "Another adult family member",
    "value": "Another adult family member",
    "id_value": "axvGUBt"
  }
]
```


---

**Node ID:** eca6cQF

**Parents:** b5nYNf6

**Children:** L5xPJtN

**Kauro:** Great!

**Kauro:** You can complete the consent process for **yourself** today in this chat

---

**Node ID:** L5xPJtN

**Parents:** eca6cQF

**Children:** Ey8gJrc

**User Transcript:** Okay, thanks

---

**Node ID:** axvGUBt

**Parents:** b5nYNf6

**Children:** mBdVTa8, 8dehgq7

**Kauro:** If your child’s other parent or another adult family member is considering enrolling, they should also complete this consent chat themselves

**Kauro:** If they have not already received a link to this consent chat, I can send them one now with just a few pieces of information

---

**Node ID:** mBdVTa8

**Parents:** axvGUBt

**Children:** AXCfXjh

**User Transcript:** They have already received a link to this chat

---

**Node ID:** AXCfXjh

**Parents:** mBdVTa8

**Children:** WnXoffX

**Kauro:** Okay, great

---

**Node ID:** 8dehgq7

**Parents:** axvGUBt

**Children:** aaVu2Vp

**User Transcript:** I’d like to send them a link to the chat

---

**Node ID:** aaVu2Vp

**Parents:** 8dehgq7

**Children:** gWJxSfh

**Kauro:** Okay!

**Kauro:** Can you tell me their name and cell phone number and/or email address?

---

**Node ID:** gWJxSfh

**Parents:** aaVu2Vp

**Children:** UiJSrG3

**Form ID:** contact-other-adult-form

**From Text:**

**From Options:**


```
[
  {
    "name": "firstname",
    "type": "text",
    "label": "First name"
  },
  {
    "name": "lastname",
    "type": "text",
    "label": "Last name"
  },
  {
    "name": "phone",
    "type": "tel",
    "label": "Phone",
    "pattern": "[0-9]{3}-[0-9]{3}-[0-9]{4}",
    "placeholder": "555-867-5309"
  },
  {
    "name": "email",
    "type": "email",
    "label": "Email",
    "placeholder": ""
  }
]
```


---

**Node ID:** UiJSrG3

**Parents:** gWJxSfh

**Children:** 4iXP7YP

**Kauro:** Thank you! We’ll contact them with the information you’ve provided

---

**Node ID:** 4iXP7YP

**Parents:** UiJSrG3

**Children:** WnXoffX

**User Transcript:** Okay, thanks

---

**Node ID:** RMb2hrx

**Parents:** b5nYNf6

**Children:** VDz6Yx8, 5ZrtE33

**Kauro:** How many of your children are you considering enrolling?

---

#### Test User Understanding

**Node ID:** VWzWNAK

**Parents:** bmorWZo, G3EXSPz

**Children:** c3jUXWd, 6PJ8q9D

**Kauro: Question 1: Are you required to participate in the PMGRC study?**

---

**Node ID:** c3jUXWd

**Parents:** VWzWNAK

**Children:** YxYN9RX

**User Transcript:** No - it's voluntary

---

**Node ID:** YxYN9RX

**Parents:** c3jUXWd

**Children:** hfQYPNY

**Kauro:** That's right!

---

**Node ID:** hfQYPNY

**Parents:** YxYN9RX, Yd6C4UC

**Children:** KyvB8Qe, 9LwDBiM

**Kauro: Question 2: Is there an alternative to enrolling in the study?**

---

**Node ID:** KyvB8Qe

**Parents:** hfQYPNY

**Children:** 9nbunFh

**User Transcript:** Yes - the alternative is to not enroll

---

**Node ID:** 9nbunFh

**Parents:** KyvB8Qe

**Children:** aBLobjJ

**Kauro:** That's correct

---

**Node ID:** aBLobjJ

**Parents:** 2N84yDZ, 9nbunFh

**Children:** BZCE5J4, KmYUkXf

**Kauro: Question 3: Is there any cost to you or your insurance provider to participate in the study?**

---

**Node ID:** BZCE5J4

**Parents:** aBLobjJ

**Children:** H6z7NJ9

**User Transcript:** Yes - I must pay (or have my insurance pay) to participate

---

**Node ID:** H6z7NJ9

**Parents:** BZCE5J4

**Children:** 3fndF8Q

**Kauro:** Actually, there will be no charge to you or your insurance for this study

**Kauro:** Participation in this study is free

---

**Node ID:** 3fndF8Q

**Parents:** H6z7NJ9

**Children:** nWC8AGV

**User Transcript:** Okay, good to know

---

**Node ID:** nWC8AGV

**Parents:** 3fndF8Q, PBhQ5Mj

**Children:** EgapBwq, oXEsSu9

**Kauro: Question 4: Will you be paid for your participation?**

---

**Node ID:** EgapBwq

**Parents:** nWC8AGV

**Children:** TEL5iQi

**User Transcript:** No

---

**Node ID:** TEL5iQi

**Parents:** EgapBwq

**Children:** LFgk3AZ

**Kauro:** That’s correct

---

**Node ID:** LFgk3AZ

**Parents:** TEL5iQi, BRcszVa

**Children:** cCwcBvK, DViRS7T, enKSA2K

**Kauro: Question 5: Will you benefit from participating in this study?**

---

**Node ID:** cCwcBvK

**Parents:** LFgk3AZ

**Children:** mye4BDq

**User Transcript:** Yes, definitely

---

**Node ID:** mye4BDq

**Parents:** cCwcBvK

**Children:** YmLFWDr

**Kauro:** Actually, that's not right

**Kauro:** Though there may be some direct benefits to you, it is also possible that you may not benefit from the study

**Kauro:** For example, it is possible you may learn about a genetic result that leads to a diagnosis or a better understanding of your health

**Kauro:** But you may not get any results from this study or any direct benefit

---

**Node ID:** YmLFWDr

**Parents:** mye4BDq

**Children:** hHuaG87

**User Transcript:** Okay, I understand

---

**Node ID:** hHuaG87

**Parents:** YmLFWDr, fcsbW4h, UpyppKe

**Children:** mcjM5XL, HSHVE6v

**Kauro: Question 6: Are there any risks from participating?**

---

**Node ID:** mcjM5XL

**Parents:** hHuaG87

**Children:** PUv6WAA

**User Transcript:** Yes, there may be risks

---

**Node ID:** PUv6WAA

**Parents:** mcjM5XL

**Children:** 2Bh3yqe

**Kauro:** You're right, there could be some risks

---

**Node ID:** 2Bh3yqe

**Parents:** PUv6WAA, j4CkZ6q

**Children:** BnpCPkN, Yj34Tnd

**Kauro: Question 7: What is the purpose of the study?**

---

**Node ID:** BnpCPkN

**Parents:** 2Bh3yqe

**Children:** FfBKuvU

**User Transcript:** To get as many samples as possible

---

**Node ID:** FfBKuvU

**Parents:** BnpCPkN

**Children:** 7zpKYiG

**Kauro:** That's actually not correct

**Kauro:** The goal of the study is to discover the causes of genetic health issues and better understand the role of genetic variants

---

**Node ID:** 7zpKYiG

**Parents:** FfBKuvU

**Children:** Wi8Xzvj

**User Transcript:** Okay, I understand

---

**Node ID:** Wi8Xzvj

**Parents:** 7zpKYiG, 7YuvMHb

**Children:** b3HAamY, mh5Ta4t

**Kauro: Question 8: Will you need to provide samples and/or data?**

---

**Node ID:** b3HAamY

**Parents:** Wi8Xzvj

**Children:** XskwUVW

**User Transcript:** Yes, the study team may collect some samples and/or data

---

**Node ID:** XskwUVW

**Parents:** b3HAamY

**Children:** JLwjXJk

**Kauro:** That's correct

---

**Node ID:** JLwjXJk

**Parents:** XskwUVW, RnQfQa9

**Children:** TBaxMHp, Mc6Kfcw

**Kauro: Question 9: Will your personal identifiable data be kept confidential?**

---

**Node ID:** TBaxMHp

**Parents:** JLwjXJk

**Children:** 22qZY38

**User Transcript:** Yes, the study team will work to keep my information confidential

---

**Node ID:** 22qZY38

**Parents:** TBaxMHp

**Children:** NPkVfwh

**Kauro:** That's right

---

**Node ID:** NPkVfwh

**Parents:** 22qZY38, F276og8

**Children:** W3wVKub, DiTdvf2

**Kauro: Question 10: If you are injured as part of your study participation, what can you do?**

---

**Node ID:** W3wVKub

**Parents:** NPkVfwh

**Children:** ebyungi

**User Transcript:** Nothing, I have to figure it out myself

---

**Node ID:** ebyungi

**Parents:** W3wVKub

**Children:** eK9JBuV

**Kauro:** Actually, that's not true

**Kauro:** If you get hurt or sick because of something to do with the study, please call the Principal Investigator, Eric Vilain, at 949-824-4090

---

**Node ID:** eK9JBuV

**Parents:** ebyungi

**Children:** Qs4fMgg

**User Transcript:** Okay, now I understand

---

**Node ID:** Qs4fMgg

**Parents:** eK9JBuV, 7ttaVVT

**Children:** nFpTPVg

**Kauro:** Thanks for answering these questions!

**Kauro:** Let me get your results...

---

**Node ID:** nFpTPVg

**Parents:** Qs4fMgg

**Children:** VUipQHn, G3EXSPz, iH6N9fF

**User Transcript:** Okay 👍

---

**Node ID:** kZ6qj4C

**Parents:** G3EXSPz

**Children:** VWzWNAK

**User Transcript:** Retry the questions.

---

**Node ID:** DiTdvf2

**Parents:** NPkVfwh

**Children:** 7ttaVVT

**User Transcript:** I can contact the principal investigator

---

**Node ID:** 7ttaVVT

**Parents:** DiTdvf2

**Children:** Qs4fMgg

**Kauro:** Yes, exactly

---

**Node ID:** Mc6Kfcw

**Parents:** JLwjXJk

**Children:** 97RE52j

**User Transcript:** No, any of my information may be shared publicly

---

**Node ID:** 97RE52j

**Parents:** Mc6Kfcw

**Children:** F276og8

**Kauro:** Actually, that’s not true

**Kauro:** The study team will take care to protect the privacy and confidentiality of your study data

---

**Node ID:** F276og8

**Parents:** 97RE52j

**Children:** NPkVfwh

**User Transcript:** Okay, that’s good to know

---

**Node ID:** mh5Ta4t

**Parents:** Wi8Xzvj

**Children:** UL94XoE

**User Transcript:** No, no samples or data are ever needed

---

**Node ID:** UL94XoE

**Parents:** mh5Ta4t

**Children:** RnQfQa9

**Kauro:** That’s not quite true

**Kauro:** In fact, the study team may need to gather some samples and data about you

**Kauro:** If you enroll, our study team will be collecting information such as health history, genetic data, and clinical records for anyone who is enrolled in this study

**Kauro:** You may also be asked to provide a sample for genetic testing, including a blood sample or a cheek swab

---

**Node ID:** RnQfQa9

**Parents:** UL94XoE

**Children:** JLwjXJk

**User Transcript:** Okay, that makes sense

---

**Node ID:** Yj34Tnd

**Parents:** 2Bh3yqe

**Children:** 7YuvMHb

**User Transcript:** To discover the causes of genetic health issues and better understand the role of genetic variants

---

**Node ID:** 7YuvMHb

**Parents:** Yj34Tnd

**Children:** Wi8Xzvj

**Kauro:** Yes, that’s correct

---

**Node ID:** HSHVE6v

**Parents:** hHuaG87

**Children:** 7YDYb3e

**User Transcript:** There are no possible risks

---

**Node ID:** 7YDYb3e

**Parents:** HSHVE6v

**Children:** j4CkZ6q

**Kauro:** Actually, there are some risks of participating in the study

**Kauro:** The main risks of this study are related to sample collection, the potential for loss of confidentiality of your information, and possible psychological risks associated with genetic information

---

**Node ID:** j4CkZ6q

**Parents:** 7YDYb3e

**Children:** 2Bh3yqe

**User Transcript:** I understand

---

**Node ID:** DViRS7T

**Parents:** LFgk3AZ

**Children:** kYibBDf

**User Transcript:** No, definitely not

---

**Node ID:** kYibBDf

**Parents:** DViRS7T

**Children:** UpyppKe

**Kauro:** Actually, that’s not right

**Kauro:** Though we can’t be sure whether you will benefit from participating in the study, it is possible that you will benefit

**Kauro:** For example, it is possible you may learn about a genetic result that leads to a diagnosis or a better understanding of your health

---

**Node ID:** UpyppKe

**Parents:** kYibBDf

**Children:** hHuaG87

**User Transcript:** Okay, I understand

---

**Node ID:** enKSA2K

**Parents:** LFgk3AZ

**Children:** fcsbW4h

**User Transcript:** Maybe, but not necessarily

---

**Node ID:** fcsbW4h

**Parents:** enKSA2K

**Children:** hHuaG87

**Kauro:** Yep, that’s right

**Kauro:** Though we can’t be sure whether you will benefit from participating in the study, it is possible that you will benefit

---

**Node ID:** oXEsSu9

**Parents:** nWC8AGV

**Children:** JsvPiaf

**User Transcript:** Yes

---

**Node ID:** JsvPiaf

**Parents:** oXEsSu9

**Children:** BRcszVa

**Kauro:** Actually, that’s not correct

**Kauro:** You will not be paid for your participation in the study

---

**Node ID:** BRcszVa

**Parents:** JsvPiaf

**Children:** LFgk3AZ

**User Transcript:** Okay, now I see

---

**Node ID:** KmYUkXf

**Parents:** aBLobjJ

**Children:** PBhQ5Mj

**User Transcript:** No - participation is free

---

**Node ID:** PBhQ5Mj

**Parents:** KmYUkXf

**Children:** nWC8AGV

**Kauro:** That's right!

---

**Node ID:** 9LwDBiM

**Parents:** hfQYPNY

**Children:** JqyNDaX

**User Transcript:** No - the only option is to enroll

---

**Node ID:** JqyNDaX

**Parents:** 9LwDBiM

**Children:** 2N84yDZ

**Kauro:** Actually, that's not true

**Kauro:** You have the option to decide NOT to enroll in the study, and this will not impact the medical care you or your family receive

---

**Node ID:** 2N84yDZ

**Parents:** JqyNDaX

**Children:** aBLobjJ

**User Transcript:** I understand

---

**Node ID:** 6PJ8q9D

**Parents:** VWzWNAK

**Children:** PWjf47L

**User Transcript:** Yes - it's required

---

**Node ID:** PWjf47L

**Parents:** 6PJ8q9D

**Children:** Yd6C4UC

**Kauro:** Actually, that's not true

**Kauro:** Participation in the study is completely voluntary

---

**Node ID:** Yd6C4UC

**Parents:** PWjf47L

**Children:** hfQYPNY

**User Transcript:** Okay, I understand

---

#### Start Consent

**Node ID:** JTCaWss

**Parents:** VUipQHn

**Children:** QKNbLGv, nbkJ3g7

**User Transcript:** Okay, sounds good

---

#### End Consent

**Node ID:** AimWGCA

**Parents:** knbtYck

**Children:** 5n6QHJ7

**Form ID:** consent-form

**From Text:**

I am the study participant or I am authorized to act on behalf of the participant.

I have read this consent form or had it read to me.

I have been invited to take part in a research study. I was told why the research is being done and how long my participation in the study is expected to last.

I was told about what will happen in the study and if there are any procedures or drugs that are experimental.

I was told that taking part in this research is voluntary. I also was told that I can decide not to take part or stop being in it at any time without any penalty to me or any change to the quality of care I receive.

I was told about the risks and possible discomforts of taking part in this research study. I was also informed if there are any possible benefits to me if I am in this study.

I have been given the chance to ask questions about the study, and my questions have been answered. If I have questions later, I can ask one of the people listed in this form.

I agree to take part in this research study.

I will receive a signed copy of this Informed Consent/Parental Permission form to keep.

**From Options:**


```
[
  {
    "name": "fullname",
    "type": "input",
    "label": "Full name",
    "input_type": "text"
  },
  {
    "name": "consent",
    "type": "checkbox",
    "label": "I consent"
  }
]
```


---

#### Deactivate Session

**Node ID:** YQHvnrA

**Parents:** aXDDbh2

**Children:**

**Kauro:** Thank you for completing the consent process with me today!

**Kauro:** For security reasons, this link will be deactivated within 24 hours

---

**Node ID:** 7PxxtTh

**Parents:** nTBdMds

**Children:**

**Kauro:** No problem, thank you for completing the consent process with me today!

**Kauro:** For security reasons, this link will be deactivated within 24 hours

---

#### Child Consent

**Node ID:** eDqsGdn

**Parents:** YkoAEVe

**Children:** nm8saM4

**Kauro:** Okay, we will now collect contact information and enrollment preferences for each child, and then we'll ask for your electronic signature

---

**Node ID:** nm8saM4

**Parents:** eDqsGdn

**Children:** 33yRxHn

**User Transcript:** Okay

---

**Node ID:** 33yRxHn

**Parents:** nm8saM4

**Children:** XGa7LFM

**Kauro:** Please provide your child's information below

---

**Node ID:** XGa7LFM

**Parents:** 33yRxHn

**Children:** afCcd5B

**Form ID:** child-consent-contact-form

**From Text:**

**From Options:**


```
[
  {
    "name": "firstname",
    "type": "input",
    "label": "First name",
    "required": true,
    "input_type": "text"
  },
  {
    "name": "lastname",
    "type": "input",
    "label": "Last name",
    "required": true,
    "input_type": "text"
  },
  {
```

```
      "name": "phone",
      "type": "input",
      "label": "Phone to use for contact regarding this child",
      "pattern": "[0-9]{3}-[0-9]{3}-[0-9]{4}",
      "required": true,
      "input_type": "tel",
      "placeholder": "555-867-5309"
    },
    {
      "name": "email",
      "type": "input",
      "label": "Email to use for contact regarding this child",
      "required": true,
      "input_type": "email",
      "placeholder": ""
    },
    {
      "name": "age_group",
      "type": "select",
      "label": "How old is this child?",
      "options": [
        {
          "label": "6 or younger",
          "value": "<=6"
        },
        {
          "label": "7 to 17",
          "value": "7-17"
        },
        {
          "label": "18+ under my guardianship",
          "value": ">=18 guardianship"
        }
      ],
      "required": true
    }
  ]
```


---

**Node ID:** afCcd5B

**Parents:** XGa7LFM

**Children:** SFiuhW9

**Kauro:** We would like to store your child’s samples (such as genetic material including DNA, RNA, or protein, or cells) in a tissue bank for future research as identified below

**Kauro:** The biobank is maintained by the study team at UCI

**Kauro:** Please let us know how you would like your child’s samples to be used

---

**Node ID:** SFiuhW9

**Parents:** afCcd5B

**Children:** Ycq8NGN

**Form ID:** sample-storage-use-form

**From Text:**

Can we store your child’s samples for purposes of this research study? (Required for participation in the study)

Can we store your child’s samples in UCI’s biobank for purposes of unrelated research?

**From Options:**


```
[
  {
    "name": "storeSamplesThisStudy",
    "type": "radio",
    "label": "Study Sample Storage",
    "options": [
      {
        "label": "Yes",
        "value": "yes"
      }
    ]
  },
  {
    "name": "storeSamplesOtherStudies",
    "type": "radio",
    "label": "Other Studies Sample Storage",
    "options": [
      {
        "label": "Yes",
        "value": "yes"
      },
      {
        "label": "No",
        "value": "no"
      }
    ]
  }
]
```


---

**Node ID:** Ycq8NGN

**Parents:** SFiuhW9

**Children:** 6KTYnrc

**Kauro:** You should know that you may change your mind at a later time and request that your sample be destroyed

**Kauro:** If you change your mind and want to request that your sample be destroyed, you must send a written request to:

Eric Vilain, MD, PhD
University of California, Irvine (UCI)
Department of Pediatrics
1003 Health Sciences Rd, Suite 308
Irvine, CA 92617

---

**Node ID:** 6KTYnrc

**Parents:** Ycq8NGN

**Children:** hbBDr5V

**User Transcript:** That's good to know

---

**Node ID:** hbBDr5V

**Parents:** 6KTYnrc

**Children:** VaSzySv

**Kauro:** We would like to store personal health information collected in this study (such as your child's name, date of birth, contact information, etc.) in a database for future research

**Kauro:** The database is maintained by the study team at UCI

**Kauro:** Please let us know how you would like your child's PHI to be stored and used

---

**Node ID:** VaSzySv

**Parents:** hbBDr5V

**Children:** VfXY5LK

**Form ID:** phi-use-form

**From Text:**

Can we store your PHI in our database for research related to this study? (Required for participation in the study)

Can we use your stored PHI to contact you about future research opportunities that are not related to this study?

**From Options:**


```
[
  {
    "name": "storePhiThisStudy",
    "type": "radio",
    "label": "PHI Storage for Study",
```

```
    "options": [
      {
        "label": "Yes",
        "value": "yes"
      }
    ]
  },
  {
    "name": "storePhiOtherStudies",
    "type": "radio",
    "label": "PHI Use for Other Studies",
    "options": [
      {
        "label": "Yes",
        "value": "yes"
      },
      {
        "label": "No",
        "value": "no"
      }
    ]
  }
]
```


---

**Node ID:** VfXY5LK

**Parents:** VaSzySv

**Children:** ZaZ73e4, J5LzZD4

**Kauro:** You may recall that we discussed the types of genetic research results that may be found in this study

**Kauro:** You get to decide which types of results you want to learn about

**Kauro:** Do you remember the different types of results we discussed, like “primary results” and “medically actionable secondary results”?

---

**Node ID:** ZaZ73e4

**Parents:** VfXY5LK

**Children:** 3Y8xK6c

**User Transcript:** Yes, I remember

---

**Node ID:** 3Y8xK6c

**Parents:** KByDnk2, ZaZ73e4

**Children:** nU9xybq

**Kauro:** Please complete the form below to let us know what types of results you would like to receive for your child

---

**Node ID:** nU9xybq

**Parents:** 3Y8xK6c

**Children:** ggfpZho

**Form ID:** result-return-form

**From Text:**

**From Options:**


```
[
  {
    "name": "rorPrimary",
    "type": "radio",
    "label": "Do you want to learn about primary results that are found?",
    "options": [
      {
        "label": "Yes",
        "value": "yes"
      },
      {
        "label": "No",
        "value": "no"
      }
    ]
  },
  {
    "name": "rorSecondary",
    "type": "radio",
    "label": "Do you also want to learn about medically actionable secondary
results?",
    "options": [
      {
        "label": "Yes",
        "value": "yes"
      },
      {
        "label": "No",
        "value": "no"
      }
    ]
  },
  {
    "name": "rorSecondaryNot",
    "type": "radio",
    "label": "Do you want to learn about secondary results that are NOT
medically actionable?",
    "options": [
```

```
      {
        "label": "Yes",
        "value": "yes"
      },
      {
        "label": "No",
        "value": "no"
      }
    ]
  }
]
```


---

**Node ID:** ggfpZho

**Parents:** nU9xybq

**Children:** nAaeApX

**Kauro:** Great! You have now entered all of your choices for your child’s enrollment

**Kauro:** You are ready to provide your signature to give consent for your child!

---

**Node ID:** nAaeApX

**Parents:** ggfpZho

**Children:** Mzke78A, Mzh9KYH

**Form ID:** consent-form

**From Text:**

I am the study participant or I am authorized to act on behalf of the participant.

I have read this consent form or had it read to me.

I have been invited to take part in a research study. I was told why the research is being done and how long my participation in the study is expected to last.

I was told about what will happen in the study and if there are any procedures or drugs that are experimental.

I was told that taking part in this research is voluntary. I also was told that I can decide not to take part or stop being in it at any time without any penalty to me or any change to the quality of care I receive.

I was told about the risks and possible discomforts of taking part in this research study. I was also informed if there are any possible benefits to me if I am in this study.

I have been given the chance to ask questions about the study, and my questions have been answered. If I have questions later, I can ask one of the people listed in this form.

I agree to take part in this research study.

I will receive a signed copy of this Informed Consent/Parental Permission form to keep.

**From Options:**

```
[
  {
    "name": "childname",
    "type": "input",
    "label": "Child\u2019s full name",
    "required": true,
    "input_type": "text"
  },
  {
    "name": "fullname",
    "type": "input",
    "label": "Full name of consenting individual (parent/legal guardian)",
    "required": true,
    "input_type": "text"
  },
  {
    "name": "consent",
    "type": "checkbox",
    "label": "I consent"
  }
]
```


---

**Node ID:** J5LzZD4

**Parents:** VfXY5LK

**Children:** 9VfACid

**User Transcript:** No, can you remind me?

---

**Node ID:** 9VfACid

**Parents:** J5LzZD4

**Children:** eXaerup

**Kauro: Primary findings** are genetic changes that may be related to the specific health concern that has been seen in your child or your family members. Primary findings often allow the clinical team to make a diagnosis

**Kauro:** Sometimes the study may find genetic variants that are not related to a patient's current known health concern

**Kauro:** These are called **secondary findings**. Some secondary findings are considered **medically actionable**, meaning if these genetic changes are known, then you or your healthcare provider can then take action to try to reduce your child's risk of developing symptoms or to better treat or manage the health concern related to the secondary findings

---

**Node ID:** eXaerup

**Parents:** 9VfACid

**Children:** gedo5CN

**User Transcript:** What kind of findings are medically actionable?

---

**Node ID:** gedo5CN

**Parents:** eXaerup

**Children:** KByDnk2

**Kauro:** Some examples of medically actionable secondary findings may be genetic variants that cause increased risk for future health concerns like heart disease or cancer

**Kauro:** Secondary findings are often related to adult-onset health risks. However, it is possible that we could discover a secondary finding that relates to symptoms in childhood

---

**Node ID:** KByDnk2

**Parents:** gedo5CN

**Children:** 3Y8xK6c

**User Transcript:** Okay

---

#### Follow Up

**Node ID:** DdxFUae

**Parents:** S2nH9zZ

**Children:** nqDwZrd

**User Transcript:** I'd prefer to talk to someone from the research team

---

**Node ID:** X4CvZHZ

**Parents:** iH6N9fF

**Children:** PZYk7Ga

**User Transcript:** Okay

---

**Node ID:** NsfXTD7

**Parents:** Dq4hG9k

**Children:** P8C5vVo

**User Transcript:** I'm not ready to decide yet

---

**Node ID:** BMeYncH

**Parents:** Dq4hG9k

**Children:** h9rpY7K

**User Transcript:** I have more questions before I decide

---

#### Decline Consent

**Node ID:** FVdUisi

**Parents:** Dq4hG9k, h9rpY7K, ixSPowJ

**Children:** Swxbt6c

**User Transcript:** I do not want to consent

---

#### Child Age Form

**Node ID:** GkgFmXm

**Parents:** 3jPsush

**Children:** mUpoZUX, gHm22vz, nGiGpjt

---

**Node ID:** mUpoZUX

**Parents:** GkgFmXm

**Children:** fmNaDgk

**Kauro: For your child who is under age 7 years**, you will have the option to complete their consent and enrollment during our chat today.

---

**Node ID:** fmNaDgk

**Parents:** mUpoZUX

**Children:** Sj26JXx

**User Transcript:** Great, thanks!

---

**Node ID:** gHm22vz

**Parents:** GkgFmXm

**Children:** gUoc5B6

**Kauro: For your child who is between age 7 and age 17**, you will have the option to give your consent for your child to enroll in the study

**Kauro:** However, because your child is over age 6, we may also need to talk to them briefly to tell them about the study. This is called assent

**Kauro:** If your child is developmentally not able to communicate, you can let us know when you talk with the team

---

**Node ID:** gUoc5B6

**Parents:** gHm22vz

**Children:** eWd55kR

**User Transcript:** Okay

---

**Node ID:** eWd55kR

**Parents:** gUoc5B6

**Children:** 5hX84fN

**Kauro:** So, after you complete the consent chat with me, I can help arrange a time for you (and your child) to talk to a member of our study team to finalize your child's enrollment.

---

**Node ID:** 5hX84fN

**Parents:** eWd55kR

**Children:** Sj26JXx

**User Transcript:** That sounds good, thanks

---

**Node ID:** nGiGpjt

**Parents:** GkgFmXm

**Children:** FMC5dJn, PEgQPMn

**Kauro:** Is **your 18+ child** still under your legal guardianship?

---

**Node ID:** FMC5dJn

**Parents:** nGiGpjt

**Children:** AoYLgFK

**User Transcript:** Yes

---

**Node ID:** AoYLgFK

**Parents:** FMC5dJn

**Children:** KgLJBEb

**Kauro:** Okay, after our chat today, you will have the option to give your consent for your child to enroll in the study.

**Kauro:** But because your child is over age 18, we may also need to speak with them briefly to tell them about the study.

---

**Node ID:** KgLJBEb

**Parents:** AoYLgFK

**Children:** k5zGiMq

**User Transcript:** Okay

---

**Node ID:** k5zGiMq

**Parents:** KgLJBEb

**Children:** DuLoHj9

**Kauro:** So, after you complete the consent chat with me, I can help arrange a time for you (and your child) to talk to a member of our study team to finalize your child's participation.

**Kauro:** If your child is developmentally not able to communicate, you can let us know when you talk with our study team.

---

**Node ID:** DuLoHj9

**Parents:** k5zGiMq

**Children:** Sj26JXx

**User Transcript:** Okay, thanks

---

**Node ID:** PEgQPMn

**Parents:** nGiGpjt

**Children:** dFVXKxp

**User Transcript:** No

---

**Node ID:** dFVXKxp

**Parents:** PEgQPMn

**Children:** eF87SCm, 3ENuxt7

**Kauro:** Individuals who are 18 years or older need to complete this consent chat themselves.

**Kauro:** Does your 18+ child already have a unique link to this chat?

---

**Node ID:** eF87SCm

**Parents:** dFVXKxp

**Children:** nj5w3by

**User Transcript:** Yes

---

**Node ID:** nj5w3by

**Parents:** eF87SCm

**Children:** WtmwJYx

**Kauro:** Okay, then they will be able to use that link to complete the consent chat themselves.

---

**Node ID:** WtmwJYx

**Parents:** nj5w3by

**Children:** Sj26JXx

**User Transcript:** Okay, thanks for letting me know

---

**Node ID:** 3ENuxt7

**Parents:** dFVXKxp

**Children:** 2b6U7Vy

**User Transcript:** No, can we send them a link?

---

**Node ID:** 2b6U7Vy

**Parents:** 3ENuxt7

**Children:** 8A8kAJS, ZykT2RX

**Kauro:** Sure!

**Kauro:** Can you tell me your child’s name, cell phone number, and/or email?

---

**Node ID:** 8A8kAJS

**Parents:** 2b6U7Vy

**Children:** ZykT2RX

**Form ID:** contact-other-adult-form

**From Text:**

**From Options:**


```
[
  {
    "name": "firstname",
    "type": "input",
    "label": "First name",
    "input_type": "text"
  },
  {
    "name": "lastname",
    "type": "input",
    "label": "Last name",
    "input_type": "text"
  },
  {
    "name": "phone",
    "type": "input",
    "label": "Phone",
    "pattern": "[0-9]{3}-[0-9]{3}-[0-9]{4}",
    "input_type": "tel",
    "placeholder": "555-867-5309"
  },
  {
    "name": "email",
    "type": "input",
    "label": "Email",
    "input_type": "email",
    "placeholder": ""
  }
```

]

---

**Node ID:** ZykT2RX

**Parents:** 2b6U7Vy, 8A8kAJS

**Children:** mGbcBU9

**Kauro:** Thank you! We'll contact them with the information you've provided.

---

**Node ID:** mGbcBU9

**Parents:** ZykT2RX

**Children:** Sj26JXx

**User Transcript:** Great, thanks!

---
